# Extracellular vesicle surface markers inform on COPD severity and mortality in COSYCONET

**DOI:** 10.64898/2026.06.30.26356923

**Authors:** Roman Martin, Katrin Laakmann, Hendrik Pott, Wilhelm Bertrams, Lennard Hinz, Isabell Burhorst, Robert Bals, Christian Herr, Anna Lena Jung, Peter Alter, Claus Franz Vogelmeier, Gernot Rohde, Bernd Schmeck, Dominik Heider

## Abstract

**Background:** Chronic obstructive pulmonary disease (COPD) is a leading cause of global morbidity and mortality, and its heterogeneity demands better biomarkers of severity and progression risk. Extracellular vesicles (EVs) are promising blood-based biomarkers, but have not been examined for COPD severity and outcomes in a large multicentre cohort.

**Methods:** We analysed 600 COSYCONET participants (up to 54 months of follow-up). EV surface markers were profiled with the MACSPlex EV Kit IO. Cross-sectional associations with severity (GOLD, FEV_1_) were primary (ordinal and linear regression); longitudinal trajectories and all-cause mortality were prespecified exploratory endpoints.

**Results:** Six EV markers showed robust associations with cross-sectional severity: CD29, CD49e and CD31 increased with severity (a cell-adhesion/matrix-remodelling signal), whereas CD81 and CD8 decreased; HLA-ABC (increasing) was less specific. No marker was linked to FEV_1_ decline. After FDR correction, lower levels of three markers with higher 54-month mortality (all HR<1): CD25 (HR 0.77, 95% CI 0.65–0.90, *q*=0.018), CD56 (HR 0.75, 95% CI 0.63–0.89, *q*=0.018) and CD142 (HR 0.74, 95% CI 0.60–0.90, *q*=0.024). CD25 and CD142 also improved reclassification, CD56 did not; a CD25 + CD69 combination showed the largest incremental signal (ΔC 0.017, 95% CI 0.002–0.032, *p*=0.027).

**Conclusion:** Circulating EV surface markers are associated with cross-sectional COPD severity. Exploratory analyses nominate CD25, CD142 and CD25 + CD69 as candidate prognostic markers requiring external validation, suggesting minimally invasive EV profiling could complement clinical assessment in COPD.

## 1. Introduction

Chronic obstructive pulmonary disease (COPD) is the third leading cause of death worldwide and affected an estimated 392 million people in 2019 (1). It is characterised by chronic airway inflammation and destruction of lung tissue, resulting in persistent, often progressive airflow limitation (2,3), and patients suffer recurrent exacerbations that drive hospitalisation and mortality (2). The burden extends beyond the lung: COPD is frequently accompanied by cardiovascular and metabolic comorbidities that compound disease severity (4–6), and current forecasts project over $24 trillion in direct and $15 trillion in indirect costs, alongside 15.60 billion exacerbations globally, by 2050 (7). Yet this growing and increasingly costly burden is borne by a strikingly heterogeneous patient population, with substantial variability in symptom burden, lung-function trajectories and disease progression (8–10). Established clinical parameters capture this variability only incompletely, and there is therefore continuing interest in biomarkers that could complement them in assessing disease activity, severity and prognosis, and ultimately improve patient care (11–13).

Extracellular vesicles (EVs) are small spherical membrane structures released by most cell types and detectable in diverse body fluids such as plasma. Thereby, EVs emerge as promising biomarkers with applications to a wide range of diseases as they can reflect the (patho)physiological state of its cell of origin (14). EVs can be classified according to their size and biogenesis pathway but also based on their protein content such as the surface expression of multi-pass transmembrane proteins like the tetraspanins CD9, CD63 and CD81, particularly in small EVs (<200 nm) (15). Additionally, EVs carry proteins and nucleic acids reflective of their cellular origin and physiological state and participate in biological processes such as immune regulation, inflammation, coagulation and intercellular communication (14,15). As EV composition may mirror systemic pathological processes and can be assessed through minimally invasive “liquid biopsies”, EVs have emerged as promising biomarkers in respiratory diseases (14,16,17). However, no large multicentre cohort has yet evaluated whether circulating EV surface markers reflect COPD severity and prognosis.

Here, we report comprehensive profiling of circulating EV surface markers in 600 participants from the multicentre COSYCONET cohort, encompassing individuals at risk for COPD (GOLD 0) as well as patients with established COPD (GOLD 1–4). We evaluated 22 EV-associated surface markers for their associations with cross-sectional disease severity, longitudinal lung function trajectories and mortality. By including GOLD 0 individuals, we were able to examine EV marker patterns across the full disease spectrum, from early at-risk states to advanced disease. Our primary aim was to identify candidate EV markers associated with cross-sectional COPD severity. Longitudinal and survival analyses were prespecified as exploratory secondary endpoints, with all prognostic findings interpreted as hypothesis-generating and requiring external validation before any clinical application.

## 2. Materials and Methods

### 2.1 Study design and population

600 participants from COSYCONET were classified across GOLD stages used in the cohort, including a subgroup categorised as GOLD 0, denoting symptomatic individuals (chronic bronchitis diagnosis, or CAT cough/phlegm score ≥3) with a post-bronchodilator FEV_1_/FVC >0.70, representing an at-risk, non-obstructed group. Sensitivity analyses restricted to GOLD 1–4 confirmed the robustness of findings. The cohort is described in **Table 1**.

**Table 1:** Baseline cohort characteristics. Mean ± SD or n (%). Baseline: all participants with EV profiling (n=600). GOLD 1–4: excludes GOLD 0 (n=547). Survival: complete cases for core Cox covariates (n=567). Per-analysis sample sizes: Table S1. “-”: not applicable by cohort definition. FEV_1_, forced expiratory volume in 1s; FEV_1_%pred, FEV_1_ percent predicted; FVC, forced vital capacity; IL-6/8, interleukin-6/8; LAMA, long-acting muscarinic antagonist; LABA, long-acting β_2_-agonist; ICS, inhaled corticosteroid; PaCO_2_, arterial CO_2_ partial pressure.

| Data | Baseline | GOLD 1–4 | Survival cohort |
| --- | --- | --- | --- |
| **Patients** | 600 | 547 | 567 |
| **Age** | 65.03 ± 8.78 | 64.71 ± 8.79 | 64.66 ± 8.86 |
| **BMI (kg/m<sup>2</sup>)** | 26.41 ± 4.75 | 26.18 ± 4.60 | 26.45 ± 4.76 |
| **Weight (kg)** | 76.91 ± 16.56 | 76.34 ± 16.45 | 76.91 ± 16.61 |
| **Height (m)** | 1.70 ± 0.09 | 1.70 ± 0.09 | 1.70 ± 0.09 |
| **Male** | 339 (56.50) | 310 (56.67) | 317 (55.91) |
| **Pack years** | 46.84 ± 38.16 | 47.63 ± 38.57 | 47.55 ± 38.46 |
| **Years with COPD** | 7.78 ± 6.84 | 7.71 ± 6.50 | 7.67 ± 6.54 |
| **GOLD 0** | 53 (8.83) | - | 48 (8.47) |
| **GOLD I** | 71 (11.83) | 71 (12.98) | 69 (12.17) |
| **GOLD II** | 232 (38.67) | 232 (42.41) | 221 (38.98) |
| **GOLD III** | 186 (31.00) | 186 (34.00) | 174 (30.69) |
| **GOLD IV** | 58 (9.67) | 58 (10.60) | 55 (9.70) |
| **FEV<sub>1</sub> (l)** | 1.66 ± 0.71 | 1.61 ± 0.69 | 1.66 ± 0.71 |
| **FEV<sub>1</sub>%pred** | 56.88 ± 20.56 | 54.85 ± 19.83 | 56.95 ± 20.50 |
| **FVC** | 3.05 ± 0.99 | 3.07 ± 0.99 | 3.07 ± 1.00 |
| **IL6 (pg/ml)** | 11.26 ± 42.57 | 11.94 ± 44.30 | 11.10 ± 43.32 |
| **IL8 (pg/ml)** | 19.26 ± 157.78 | 20.28 ± 164.48 | 19.58 ± 160.89 |
| **C-reactive protein** | 10.97 ± 28.88 | 11.09 ± 29.91 | 10.90 ± 28.77 |
| **any LAMA** | 436 (72.67) | 413 (75.50) | 418 (73.72) |
| **any LABA** | 490 (81.67) | 455 (83.18) | 462 (81.48) |
| **any ICS** | 385 (64.17) | 360 (65.81) | 362 (63.84) |
| **Leukocyte** | 7.89 ± 2.36 | 7.95 ± 2.36 | 7.86 ± 2.36 |
| **Platelet** | 258.57 ± 70.96 | 261.23 ± 72.05 | 258.68 ± 70.89 |
| **PaCO<sub>2</sub>** | 38.22 ± 4.88 | 38.19 ± 4.90 | 38.20 ± 4.87 |

To assess the clinical relevance of EV surface markers in COPD, we applied a structured analytical framework examining cross-sectional disease severity, longitudinal lung function decline and survival outcomes (see **Figure 1**).

**Figure 1.**
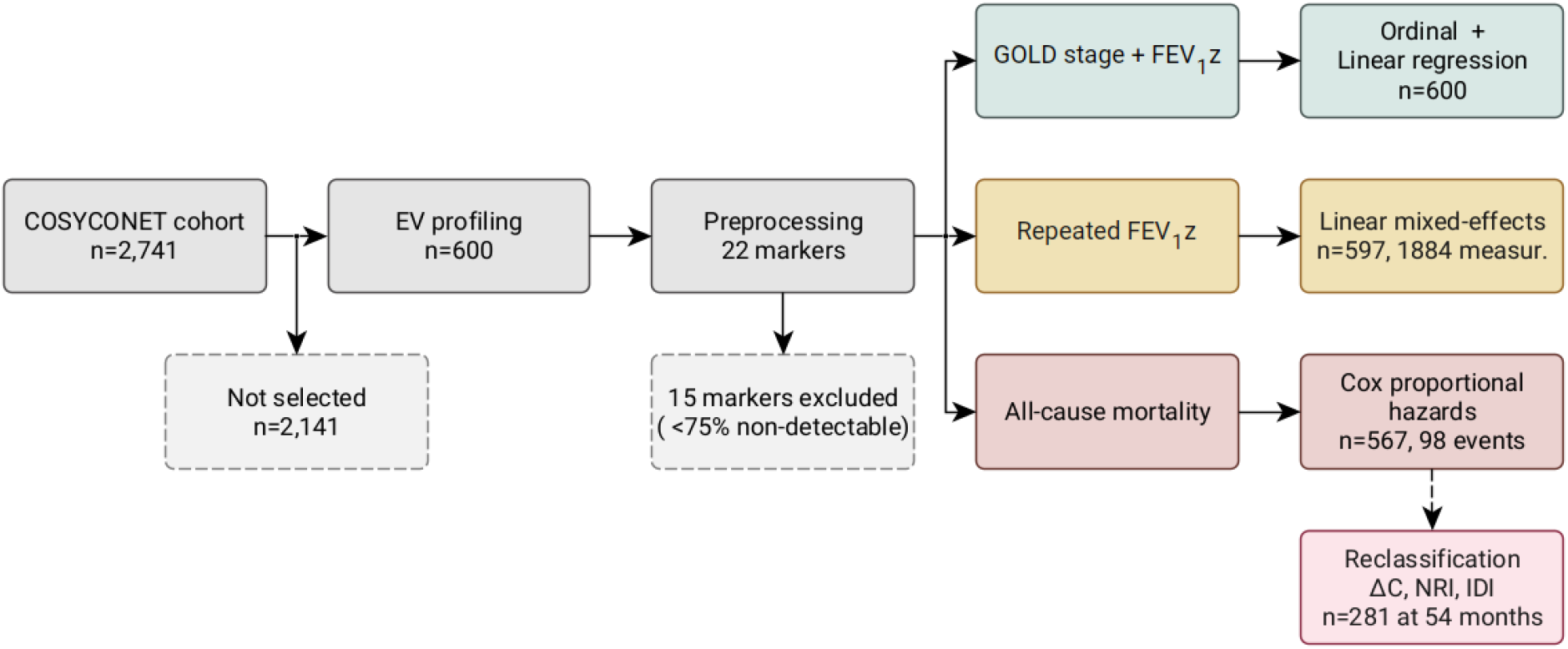
Study design and analytical framework. Participants flow from the COSYCONET cohort (n=2,741) through EV profiling (n=600) to the three analytical endpoints. Fifteen of 37 markers were excluded for low detectability (detectable in fewer than 75% of samples), leaving 22 for analysis. Reclassification analyses were restricted to n=281 participants with known event status at the 54-month horizon. The three analytical arms: cross-sectional severity (ordinal and linear regression), longitudinal lung function (linear mixed-effects models), and survival (Cox regression), with incremental prognostic evaluation (ΔC-index, NRI, IDI) downstream of the Cox analyses. Covariate sets per analysis: **Table S1**.

### 2.2 Blood sampling and surface marker profiling

Peripheral blood was collected at enrolment in BD™ P100 tubes (BD Biosciences), centrifuged within one hour of collection and plasma was stored at -80°C. EV surface markers were profiled by flow cytometry using the MACSPlex EV kit IO (Miltenyi Biotec) on ultracentrifuged plasma (100,000 × g, 1 h, 4°C; Sorvall Discovery 90SE; Thermo Fisher Scientific; 70Ti rotor, Beckman Coulter, USA in X100 PA ultracentrifuge microtubes, Thermo Fisher Scientific) and further processed according to the manufacturer’s protocol. Detailed protocols for sample handling, ultracentrifugation, staining, and acquisition are provided in the Supplementary Methods.

### 2.3 Surface Marker Preprocessing

Markers detected in fewer than 75% of cohort samples were excluded (**Figure S1, Table S2**), leaving 22 markers for analysis. Intensities were log1p-transformed, Winsorised at the 1st and 99th percentiles, z-scored (**Figure S2**), and batch-corrected using ComBat (18,19). Full preprocessing parameters are detailed in Supplementary Methods.

Analyses addressed cross-sectional severity, longitudinal FEV_1_ trajectories, and mortality. The clinical baseline + FEV_1_z covariate set served as the primary adjustment; alternative lung function adjustments were sensitivity analyses (Table S1). Benjamini-Hochberg FDR correction was applied to all p-values from primary analyses across the 22 markers. FEV_1_ and FVC z-scores used the GLI reference equations (20); RV/TLC z-scores from Hall (21). In sensitivity analyses, results were consistent with the multi-ethnic reference (22). Z-scores were computed with pyspiro (23).

#### 2.3.1 Ordinal logistic and linear regression for disease severity

Covariates were selected a priori from LASSO screening combined with clinical domain expertise. GOLD stage was modelled using proportional-odds ordinal logistic regression; odds ratios are reported with 95% CIs and McFadden’s pseudo-R^2^. Incremental marker contribution was assessed using likelihood-ratio tests. As supportive analyses, two-sided Jonckheere-Terpstra tests were performed to confirm monotonic trends across GOLD stages.

Associations with continuous lung function were assessed using linear regression models with FEV_1_ z-scores as the dependent variable. Ordinary least squares models were fitted with heteroscedasticity-robust standard errors (HC3). Incremental explanatory value of each marker was quantified using partial R^2^.

#### 2.3.2 Longitudinal analyses

Longitudinal FEV_1_ z-score trajectories were modelled by linear mixed-effects regression with fixed effects for the standardised EV marker, follow-up time (years from baseline, centred at the cohort mean), their interaction, and the disease-severity covariate set (sex, age, BMI, smoking state, pack-years, years with COPD, SGRQ activity); subject-specific random intercepts and random slopes for time were included. IL-6, included in the cross-sectional models, was excluded here because its inclusion caused the mixed-effects model to fail at fitting; the remaining covariates were held identical to the cross-sectional analyses. Per-analysis covariate sets are summarised in Table S1.

#### 2.3.3 Survival analyses

Multivariable associations with all-cause mortality were assessed using Cox proportional hazards regression with Efron handling of tied event times and robust sandwich standard errors. The proportional hazards assumption was checked using scaled Schoenfeld residuals (**Figure S5**).

Incremental prognostic information of individual EV markers was assessed by comparing the clinical Cox model with an otherwise identical model additionally including one candidate marker. Discrimination was quantified using Harrell’s C-index; reclassification at the prespecified 54-month horizon was quantified using continuous and categorical NRI (cut-points 5%, 10%, 20%) and IDI. Ninety-five per cent confidence intervals and two-sided p-values were obtained by bootstrap resampling (5000 resamples; **Figure S7**). Reclassification analyses were restricted to participants with known event status at the horizon. Calibration is shown in **Figure S6**.

A combined-marker analysis was conducted as a predefined exploratory step. The five EV markers with the highest mean outer-fold selection frequency in repeated nested cross-validation (5×5 folds, 10 repeats; all ≥0.92) were retained (**Table S9a**), and all ten pairs were ranked by bootstrap ΔC-index against the clinical backbone (**Table S9b**). The same five markers also showed the largest absolute standardised coefficients in a ridge-penalised Cox model on the full feature set, confirming the shortlist by an independent ranking. Optimism in the incremental prognostic estimates was assessed by repeated stratified 10-fold cross-validation (100 repeats; **Table S9c**). Full procedural details for the nested CV, pair search, and optimism correction are provided in Supplementary Methods.

## 3. Results

### 3.1 EV marker associations with COPD severity

We screened all 22 EV surface markers for associations with COPD severity using ordinal logistic regression for GOLD stage and linear regression for FEV_1_. In the primary analyses, nine overlapping candidate biomarkers were significant in both models after FDR correction (q<0.05): CD8, CD9, CD29, CD31, CD49e, CD69, CD81, CD146, and HLA-ABC. Direction was consistent across both models: CD29, CD49e, CD31 and HLA-ABC increased with higher GOLD stage and lower FEV_1_, whereas CD81 and CD8 were higher in milder disease.

Across all sensitivity analyses (GOLD 1–4 only, no covariates, additional adjustment for FVC or RV/TLC), six markers remained robust, defined as q<0.01 in both main models and p<0.05 in every sensitivity analysis with a consistent effect direction throughout: CD29 (OR 1.526, β −0.168), CD49e (OR 1.352, β −0.147), CD31 (OR 1.380, β −0.141), CD81 (OR 0.696, β 0.162), HLA-ABC (OR 1.497, β −0.147) and CD8 (OR 0.747, β 0.131). See **Table 2** and **Figure 2, Table S3** and **Figure S3** for all sensitivity analyses.

**Table 2:** EV surface markers associated with COPD severity. OR (95% CI) for higher GOLD stage from proportional-odds ordinal logistic regression; β (95% CI) for FEV_1_ z-score from linear regression with HC3-robust SE. Both per 1-SD increase in standardised marker level, adjusted for the prespecified covariate set (sex, age, BMI, smoking state, pack-years, years with COPD, SGRQ activity, IL-6; n=600). q: Benjamini-Hochberg-adjusted p across 22 markers. Full results for all 22 markers: Table S3. Markers bolded in Figure 2 (CD8, CD29, CD31, CD49e, CD81, HLA-ABC) remained significant across all sensitivity analyses (Tables S3b.2-S3b.6).

| Marker | GOLD stages |  | FEV<sub>1</sub>z |  |
| --- | --- | --- | --- | --- |
| | q | OR (95% CI) | q | $\beta$ (95% CI) |
| **CD8** | 0.0010 | 0.7470 (0.6358 to 0.8776) | 0.0029 | 0.1315 (0.0535 to 0.2094) |
| CD9 | 0.0009 | 1.3481 (1.1493 to 1.5814) | 0.0029 | -0.1353 (-0.2162 to -0.0545) |
| **CD29** | <0.0001 | 1.5261 (1.2936 to 1.8005) | 0.0004 | -0.1677 (-0.2440 to -0.0915) |
| **CD31** | 0.0005 | 1.3804 (1.1753 to 1.6213) | 0.0020 | -0.1410 (-0.2198 to -0.0622) |
| **CD49e** | 0.0009 | 1.3523 (1.1485 to 1.5922) | 0.0014 | -0.1468 (-0.2244 to -0.0693) |
| CD69 | 0.0008 | 1.3637 (1.1592 to 1.6042) | 0.0046 | -0.1269 (-0.2069 to -0.0469) |
| **CD81** | 0.0001 | 0.6958 (0.5897 to 0.8211) | 0.0014 | 0.1623 (0.0781 to 0.2466) |
| CD146 | 0.0009 | 1.3463 (1.1442 to 1.5840) | 0.0029 | -0.1383 (-0.2211 to -0.0555) |
| **HLA-ABC** | <0.0001 | 1.4971 (1.2690 to 1.7662) | 0.0014 | -0.1469 (-0.2255 to -0.0683) |

**Figure 2.**
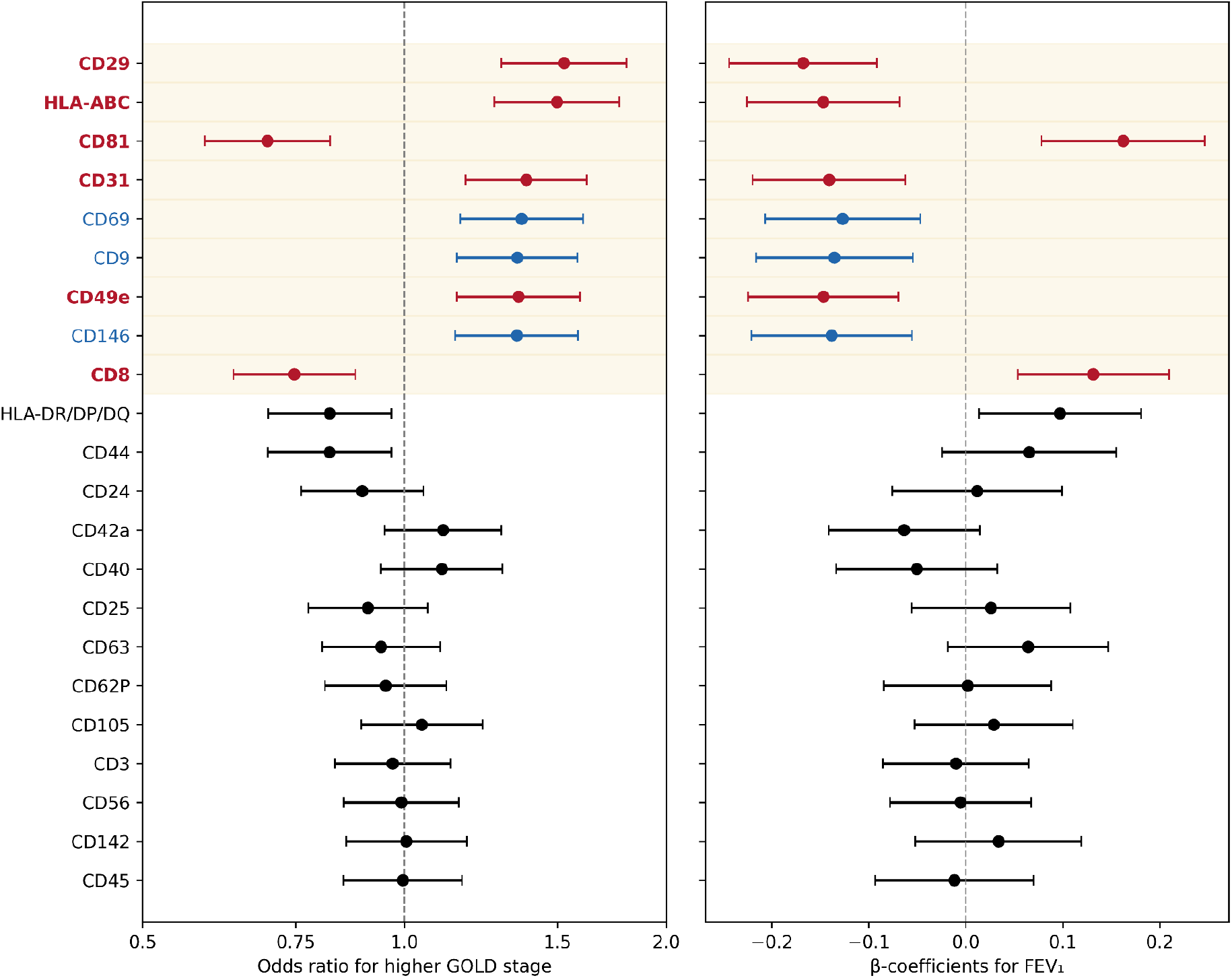
Cross-sectional associations of EV surface markers with COPD severity. Forest plots show odds ratios for higher GOLD stage (left, ordinal logistic regression) and β-coefficients for FEV_1_ z-score (right, linear regression) with 95% CI for all 22 markers. Red: markers reaching q<0.05 in both models and robust across all sensitivity analyses (CD8, CD29, CD31, CD49e, CD81, HLA-ABC). Blue: additional markers reaching q<0.05 in both models. Full numerical results are provided in Table S3.

Monotonic trends across GOLD stages were confirmed by FDR-adjusted Jonckheere-Terpstra tests in both the full cohort and the GOLD 1–4 subgroup (**Table S4**), indicating that marker levels shifted stepwise with each successive stage rather than only at the extremes.

### 3.2 Longitudinal evaluation of EV markers and lung function decline

In adjusted linear mixed-effects models of longitudinal FEV_1_ (**Table S6, Figure S4**), no marker showed a significant marker-by-time interaction after FDR correction. CD81 showed the most notable signal, a significant adjusted main effect (q<0.001) and a nominal interaction in the GOLD 1–4 subgroup (p=0.033), but the interaction term did not overcome correction in the full cohort (p=0.089, q=0.523). Unadjusted progressor/non-progressor comparisons (**Table S5**) did not persist in covariate-adjusted models. Overall, baseline EV marker levels were not robustly associated with subsequent FEV_1_ decline.

### 3.3 Survival analysis for mortality

EV surface markers were evaluated in Cox proportional hazards regression adjusted for established clinical parameters. All three associations (q<0.05) were protective in direction (HR<1), i.e. lower marker levels corresponded to higher mortality risk:: CD25 (HR 0.766, 95% CI 0.649-0.904, *q*=0.018), CD56 (HR 0.749, 95% CI 0.627-0.894, *q*=0.018) and CD142 (HR 0.740, 95% CI 0.605-0.904, *q*=0.024). All three associations were robust across alternative lung function adjustment schemes (**Table S7**).

At the prespecified 54-month horizon, the reference model (Core + FEV_1_z) achieved a C-index of 0.823 (n=567). Single markers produced modest gains in discrimination (CD25 ΔC 0.009, *p*=0.156; CD56 0.014, *p*=0.062; CD142 0.010, *p*=0.128). CD25 and CD142 significantly improved risk reclassification across all metrics (CD25: continuous NRI 0.352, *p*=0.020; categorical NRI 0.183, *p*=0.006; IDI 0.017, *p*=0.026. CD142: 0.441, *p*=0.003; 0.156, *p*=0.019; 0.020, *p*=0.015). CD56, despite the lowest p-value, reached significance on no reclassification metric (**Table 3, Figure 3, Table S8**). For CD25, the discrimination gain seen without FEV_1_z adjustment (ΔC *p*=0.040) was attenuated once FEV_1_ was included (*p*=0.156); CD56 showed an analogous pattern in its integrated discrimination (IDI significant at p=0.022 without FEV_1_z, p=0.106 with), indicating partial overlap of both signals with airflow obstruction.

**Table 3:** Incremental prognostic performance at the 54-month horizon. n=567 (98 events) for all Cox models; reclassification analyses restricted to n=281 participants with known event status at 54-month horizon. Reference model: sex, age, leukocyte, PaCO_2_, platelet, FEV_1_ z-score. 95% CI and two-sided p-values from 5000 bootstrap resamples. Categorical NRI cut-points: 5%, 10%, 20%. CD25 + CD69 identified by the predefined selection procedure. Bold: p<0.05. See Table S8 for sensitivity across lung function adjustments.

| Marker | C-index | $\Delta C$ (95% CI, p) | cont. NRI (95% CI, p) | cat. NRI (95% CI, p) | IDI (95% CI, p) |
| --- | --- | --- | --- | --- | --- |
| - | 0.823 | - | - | - | - |
| CD25 | 0.832 | 0.009 (-0.003 to 0.022, 0.156) | 0.352 (0.056-0.636, **0.020**) | 0.183 (0.058-0.311, **0.006**) | 0.017 (0.002-0.032, **0.026**) |
| CD56 | 0.837 | 0.014 (0.000-0.031, 0.062) | 0.236 (-0.061 to 0.524, 0.121) | 0.116 (-0.009 to 0.241, 0.066) | 0.013 (-0.003 to 0.030, 0.106) |
| CD142 | 0.834 | 0.010 (-0.003 to 0.025, 0.128) | 0.441 (0.159-0.723, **0.003**) | 0.156 (0.025-0.295, **0.019**) | 0.020 (0.004-0.036, **0.015**) |
| CD25+<br>CD69 | 0.840 | 0.017 (0.002-0.032, **0.027**) | 0.468 (0.182-0.734, **0.002**) | 0.183 (0.045-0.324, **0.011**) | 0.024 (0.004-0.043, **0.019**) |

**Figure 3:**
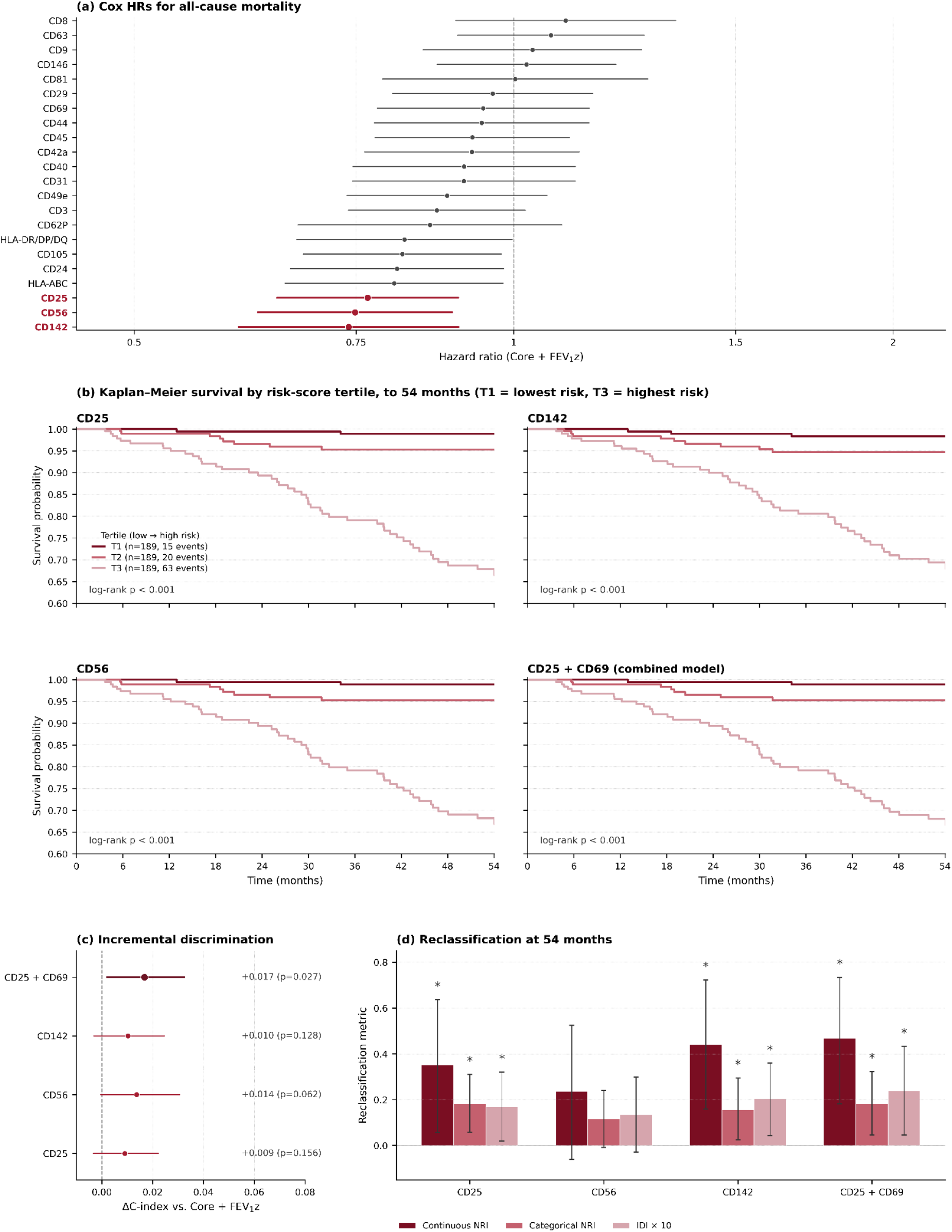
Survival findings for EV surface markers. Reference model throughout: core covariates + FEV_1_ z-score. (a) Adjusted Cox hazard ratios with 95% CI for all 22 markers; red indicates FDR-significant markers (q<0.05): CD25, CD56 and CD142. (b) Kaplan-Meier curves by Cox risk-score tertile (T1 = lowest risk, T3 = highest risk; n=189 per tertile) for CD25, CD142, CD56 and the predefined CD25 + CD69 combination, truncated at the 54-month horizon, with across-tertile log-rank p. (c) ΔC-index versus the reference model with 95% CI for CD25, CD56, CD142 and the CD25 + CD69 combination. (d) Reclassification metrics at 54 months: continuous NRI, categorical NRI (cut-points 5%, 10%, 20%) and IDI (×10), for the same four configurations. Asterisks: p<0.05 (two-sided bootstrap, 5000 resamples). Sensitivity analyses: Tables S7-S8.

In a predefined exploratory combined-marker analysis, repeated nested cross-validation identified five EV markers with the highest selection stability, confirmed by an independent ranking on standardised ridge coefficients (CD62P, CD142, CD69, HLA-ABC, CD25; **Table S9a**). Exhaustive evaluation of all ten pairs (**Table S9b**) ranked CD25 + CD69 highest by ΔC-index (ΔC 0.017, 95% CI 0.002-0.032, *p*=0.027; continuous NRI 0.468, p=0.002; categorical NRI 0.183, *p*=0.011; IDI 0.024, *p*=0.019), with improvements consistent across all seven adjustment schemes (ΔC p range 0.001-0.043). Within the joint model CD25 remained predictive (HR 0.702, 95% CI 0.584-0.845, *p*<0.001) while CD69 did not (HR 1.17, 95 % CI 0.922-1.486, *p*=0.197), consistent with complementary rather than independent information. Subgroup analyses showed no evidence of effect modification (**Figure S8**).

Cross-validated estimates (**Table S9c**) showed minimal optimism: apparent and CV-median ΔC-index, continuous NRI, categorical NRI and IDI agreed closely across all four configurations (|optimism| ≤ 0.005 for ΔC and IDI; |optimism| ≤ 0.10 for reclassification metrics). CD25 + CD69 retained the largest CV-median gains (ΔC 0.013, cNRI 0.440, catNRI 0.120, IDI 0.021).

## 4. Discussion

In this large multicentre cohort, circulating EV surface markers showed three principal patterns. First, six markers were robustly associated with cross-sectional disease severity across both the full GOLD 0–4 spectrum and the established-disease GOLD 1–4 subgroup. These resolved into a coherent cell-adhesion/matrix-remodelling axis (CD29, CD49e, CD31), all rising with disease severity, together with three less mechanistically specific signals: the near-ubiquitous EV constituents CD81 and HLA-ABC, and the cytotoxic-lymphocyte marker CD8. Notably, the direction differed across these markers: HLA-ABC increased with severity, whereas CD81 and CD8 were higher in milder disease. Second, no marker was robustly associated with longitudinal FEV_1_ decline after FDR correction. Third, in exploratory survival analyses, CD25, CD56 and CD142 were each independently associated with all-cause mortality, with all hazard ratios below unity, that is, lower circulating levels marked higher risk. Their reclassification profiles diverged: CD25 and CD142 improved all three metrics, whereas CD56, despite the lowest Cox p-value, improved none. CD25 and CD142 therefore emerge as the most promising single-marker prognostic candidates, and although CD69 alone was not associated with mortality, its combination with CD25 yielded the largest exploratory incremental signal, consistent with complementary information from two lymphocyte-activation markers.

The most mechanistically coherent severity signal is a cell-adhesion and matrix-remodelling axis. The α5β1 fibronectin receptor formed by CD49e (α5) and CD29 (β1) mediates cell-matrix adhesion and tissue remodelling (24,25), processes central to emphysematous destruction and airway-wall remodelling; the parallel associations of both subunits, both increasing with severity, point to a single receptor system rather than two independent markers. CD31^+^ (PECAM-1) EVs largely derive from platelets and endothelium and reflect the vascular activation characteristic of COPD (26,27), adding a vascular dimension to this axis.

The remaining robust associations are statistically reliable but harder to assign to a discrete pathway. CD81 is a canonical small-EV tetraspanin (28,29) and HLA class I (HLA-ABC) is constitutively expressed on virtually all nucleated cells and platelets and is a common EV constituent (15,30); for both, the signal may partly reflect overall EV abundance rather than a specific process. Notably, the two move in opposite directions with severity (HLA-ABC increasing, CD81 decreasing), which argues against a single total-vesicle-abundance confound but does not establish a specific mechanism for either. CD8, a cytotoxic-T-lymphocyte marker, was equally robust and associated with lower severity; we report it as a distinct, robust candidate rather than part of the highlighted axis.

The prognostic markers point to a different biological compartment. CD25 and CD69 are central to T-cell activation (31,32). All three mortality-associated markers had hazard ratios below unity: lower circulating levels marked higher risk. For CD25, a key regulator of T-cell function, this is biologically plausible, pointing to impaired immune regulation, though the association should not be over-interpreted mechanistically. CD25 was independently predictive, CD69 alone was not, and their combination outperformed either marker singly - consistent with complementary aspects of the systemic immune state. CD56 (NCAM-1), the canonical NK-cell marker (33), showed a strong Cox association without reclassification gain, compatible with a broad signal of preserved immune competence rather than individual risk contribution. CD142 (tissue factor) is a transmembrane receptor that initiates the extrinsic coagulation cascade upon vascular injury (34). It can be induced on monocytes and endothelial cells during inflammation (35), and procoagulant circulating EVs may contribute to the cardiovascular comorbidity burden of COPD (27). The protective direction observed here is therefore biologically counterintuitive and may reflect prevalent-cohort selection, whereby individuals with the most adverse procoagulant profiles experienced fatal events before enrolment, or residual confounding by antiplatelet and anticoagulant therapy, and warrants dedicated mechanistic validation.

That the prognostic signal (CD25, CD69, CD142) shifts towards immune activation and coagulation, a compartment distinct from the adhesion/remodelling axis associated with severity, may explain why the severity- and mortality-associated markers are largely non-overlapping. This separation should be interpreted cautiously: four of the five markers in the stability-based shortlist (**Table S9a)** carried negative coefficients, a pattern unlikely to reflect five independent protective mechanisms and more plausibly indicating shared prevalent-cohort selection. External validation in incident-case cohorts will be required to distinguish genuine protective associations from selection-driven artefacts.

For the clinician, the relevant question is whether an EV marker re-stratifies individual patients beyond established predictors. This was captured more clearly by reclassification (NRI/IDI) than by the c-index, consistent with the recognised insensitivity of the c-statistic to incremental improvement over a well-performing base model (36,37). CD25 and CD142 exemplify this, whereas CD56’s Cox significance without reclassification gain illustrates that the two measures are conceptually distinct and should not be conflated. The exploratory CD25 + CD69 combination improved both domains, suggesting that multi-marker EV profiles may be more informative than single markers, pending independent validation.

Several limitations should be considered. EV surface markers were measured at baseline only, precluding assessment of intra-individual temporal change. Although the MACSPlex EV kit offers a scalable, standardised platform for screening large cohorts, it provides semiquantitative surface-marker measurements rather than absolute vesicle counts, and the observed signals cannot be assigned with certainty to specific cellular sources, although a meaningful narrowing of candidate EV-secreting cell types remains feasible. Despite multicentre recruitment and extensive phenotyping, complete-case analyses and model-specific differences in covariate availability may have introduced selection effects, and the sample size at the 54-month horizon constrained the number of markers that could be jointly modelled. Finally, the performance estimates from combined CD25 + CD69 models in a single cohort remain susceptible to optimism (38); moreover, the NRI has been criticised as an unreliable measure of incremental value even with independent data (39,40). Both considerations remain provisional until externally replicated.

Taken together, the strongest findings were cross-sectional: circulating EV surface markers are associated with COPD severity, most coherently through a cell-adhesion/matrix-remodelling axis (CD29, CD49e, CD31). Exploratory survival analyses nominate CD25, CD142 and CD25 + CD69 as candidate prognostic markers requiring external validation. Minimally invasive EV profiling may nonetheless inform future COPD biomarker development.

## Data Availability

The data are part of the German COPD cohort COSYCONET (www.asconet.net) and are available upon reasonable request under controlled access.

## Ethics

COSYCONET was conducted in accordance with the Declaration of Helsinki and the principles of Good Clinical Practice. Ethical approval was granted by the coordinating committee in Marburg (Ethikkommission FB Medizin Marburg) together with the responsible committees at each participating site: Bad Reichenhall (Ethikkommission bayerische Landesärztekammer); Berlin (Ethikkommission Ärztekammer Berlin); Bochum (Ethikkommission Medizinische Fakultät der RUB); Borstel (Ethikkommission Universität Lübeck); Coswig (Ethikkommission TU Dresden); Donaustauf (Ethikkommission Universitätsklinikum Regensburg); Essen (Ethikkommission Medizinische Fakultät Duisburg-Essen); Gießen (Ethikkommission Fachbereich Medizin); Greifswald (Ethikkommission Universitätsmedizin Greifswald); Großhansdorf (Ethikkommission Ärztekammer Schleswig-Holstein); Hamburg (Ethikkommission Ärztekammer Hamburg); MHH Hannover/Coppenbrügge (MHH Ethikkommission); Heidelberg Thorax/Uniklinik (Ethikkommission Universität Heidelberg); Homburg (Ethikkommission Saarbrücken); Immenhausen (Ethikkommission Landesärztekammer Hessen); Kiel (Ethikkommission Christian-Albrechts-Universität zu Kiel); Leipzig (Ethikkommission Universität Leipzig); Löwenstein (Ethikkommission Landesärztekammer Baden-Württemberg); Mainz (Ethikkommission Landesärztekammer Rheinland-Pfalz); München LMU/Gauting (Ethikkommission Klinikum Universität München); Nürnberg (Ethikkommission Friedrich-Alexander-Universität Erlangen Nürnberg); Rostock (Ethikkommission Universität Rostock); Berchtesgadener Land (Ethikkommission Land Salzburg); Schmallenberg (Ethikkommission Ärztekammer Westfalen-Lippe); Solingen (Ethikkommission Universität Witten-Herdecke); Ulm (Ethikkommission Universität Ulm); and Würzburg (Ethikkommission Universität Würzburg). All participants provided written informed consent, which included consent to publish data collected over the course of the study. Recruitment within the COSYCONET framework comprised 2741 patients (ClinicalTrials.gov identifier NCT01245933; first registered 18 November 2010).

## Author contributions

**Roman Martin**: Conceptualization, Data curation, Formal analysis, Investigation, Methodology, Software, Supervision, Validation, Visualization, Writing - original draft, Writing - review & editing. **Katrin Laakmann**: Conceptualization, Data curation; Formal analysis, Methodology, Project administration, Resources, Validation, Writing - original draft, Writing - review & editing. **Hendrik Pott**: Conceptualization, Data curation, Formal analysis, Methodology, Software, Validation, Writing - original draft, Writing - review & editing. **Wilhelm Bertrams**: Conceptualization, Data curation; Formal analysis, Methodology, Project administration, Resources, Validation, Writing - original draft, Writing - review & editing. **Lennard Hinz**: Resources, Software. **Isabell Burhorst**: Investigation, Resources, Resources, Writing - original draft, Writing - review & editing. **Robert Bals**: Resources, Writing - original draft, Writing - review & editing. **Christian Herr**: Resources, Writing - original draft, Writing - review & editing. **Anna Lena Jung**: Conceptualization, Resources, Writing - original draft, Writing - review & editing. **Peter Alter**: Conceptualization, Data curation, Writing - original draft, Writing - review & editing. **Claus Franz Vogelmeier**: Conceptualization, Funding acquisition, Investigation, Project administration, Resources, Supervision, Validation, Writing - original draft, Writing - review & editing. **Gernot Rohde**: Conceptualization, Funding acquisition, Investigation, Project administration, Resources, Supervision, Validation, Writing - original draft, Writing - review & editing. **Bernd Schmeck**: Conceptualization, Funding acquisition, Investigation, Project administration, Resources, Supervision, Validation, Writing - original draft, Writing - review & editing. **Dominik Heider**: Conceptualization, Funding acquisition, Investigation, Project administration, Resources, Supervision, Validation, Writing - original draft, Writing - review & editing.

## Conflict of interest

The authors declare that they have no competing interests.

## Funding

This research was funded by the German Federal Ministry of Research, Technology and Space (BMFTR) under the project PerMed-COPD (grant number 01EK2203F) and the Hessisches Ministerium für Wissenschaft und Kunst (LOEWE Habitat).

## Acknowledgements

We thank Isabell Beinborn for excellent technical assistance.

## Notes

### Competing Interest Statement

The authors have declared no competing interest.

### Author Declarations

The COSYCONET study was approved by the coordinating ethics committee in Marburg (Ethikkommission FB Medizin Marburg) and by the responsible ethics committees at all participating study sites, and was conducted in accordance with the Declaration of Helsinki and Good Clinical Practice. All participants provided written informed consent, including consent to publish the data collected. ClinicalTrials.gov identifier NCT01245933 (first registered 18 November 2010).

