## Supplementary Information for "Extracellular vesicle surface markers inform on COPD severity and mortality in COSYCONET"

### Supplementary Methods

#### S1 Blood Sample Collection for Extracellular Vesicles

Blood samples were obtained according to the COSYCONET study protocol. Briefly, peripheral blood was collected upon enrolment from COPD patients across GOLD stages using BD™ P100 tubes containing K2EDTA anticoagulant and proprietary protease inhibitors (BD Biosciences). Tubes were gently inverted (8-10 times) and centrifuged at 2,500 × g for 15 minutes at 20°C immediately after collection. Plasma was transferred to fresh tubes and stored at -80°C within one hour until further processing.

#### S2 Surface Marker Profiling

The EV surface marker profile was analysed using the MACSPlex EV kit IO (Miltenyi Biotec). P100 Plasma samples (125 µL) from 600 COSYCONET participants were diluted with PBS to 1.5 mL and ultracentrifuged at 100,000 × g for 1 h at 4°C. The supernatant was carefully removed, and the pellet was resuspended in the kit assay buffer to reach a final volume of 130 µL/sample. 120 µL/sample was used for EV surface marker staining according to the manufacturer's protocol. Flow cytometric acquisition and analysis were performed according to the manufacturer's protocol using the BD FACSymphony™ A1 flow cytometer with the BD FACSDiva Software Version 9.0.2. and FlowJoTM (v10.10) analysis software. The relative EV marker levels (median signal intensity) were calculated as recommended by the manufacturer's protocol.

#### S3 Surface Marker Preprocessing

Median signal intensities of 0.0 were first treated as non-detectable and set to missing for prevalence assessment. Markers with detectable (non-zero) signals in fewer than 75% of cohort samples were excluded from all downstream analyses (see **Figure S1**, **Table S2**). For the 22 retained markers, non-detectable values were restored to 0.0 for statistical modelling, consistent with the convention that absence of detectable signal represents a true low value rather than missing data. All EV marker intensities were log1p-transformed to reduce right-skewness and stabilise variance. Values were subsequently mildly Winsorised at the 1st and 99th percentiles to limit the influence of extreme observations, and then standardised using z-scores to enable comparability across markers and improve numerical stability in multivariate models (see **Figure S2**). To account for potential technical variation between measurement batches, marker intensities were subsequently batch-corrected using ComBat.

#### S4 Combined marker selection and optimism correction

A broad set of clinically relevant variables, including demographic characteristics, smoking exposure, lung function and laboratory parameters, was evaluated using ridge-penalised Cox regression to derive a stable clinical backbone model, reducing coefficient variance in the presence of correlated predictors.

The combined-marker model was identified by repeated nested cross-validation using elastic-net-penalised Cox models across a 25 × 4 penalty/L1-ratio grid. Five outer folds were stratified on event status; an inner 5-fold loop selected the (penalty, L1-ratio) pair maximising the mean inner C-index. The procedure was repeated 10 times with different random seeds, and the mean outer-fold selection frequency across all 50 outer folds defined the stability score per marker. The five markers with the highest stability scores were retained as the candidate set. All ten pairs from this set were evaluated exhaustively by bootstrap ΔC-index, continuous NRI, categorical NRI and IDI (5000 resamples, 54-month horizon) against the clinical backbone.

To quantify optimism in the incremental prognostic estimates, repeated stratified 10-fold cross-validation (100 repeats, 1000 evaluations per configuration) was performed for the four primary configurations. Within each training fold, winsorisation bounds and z-scoring parameters were re-fit on training data and applied to the held-out fold; ComBat batch correction was applied once on the full dataset prior to cross-validation. The reference (Core + $FEV_1z$) and extended models were fitted on each training fold, and held-out predictions used to compute all five metrics. CV-corrected estimates were summarised as the median across the 1000 evaluations; optimism was defined as apparent - CV-median. Percentile intervals (2.5-97.5%) reflect fold-to-fold variability and are reported as a stability diagnostic, not as confidence intervals.

### Figures

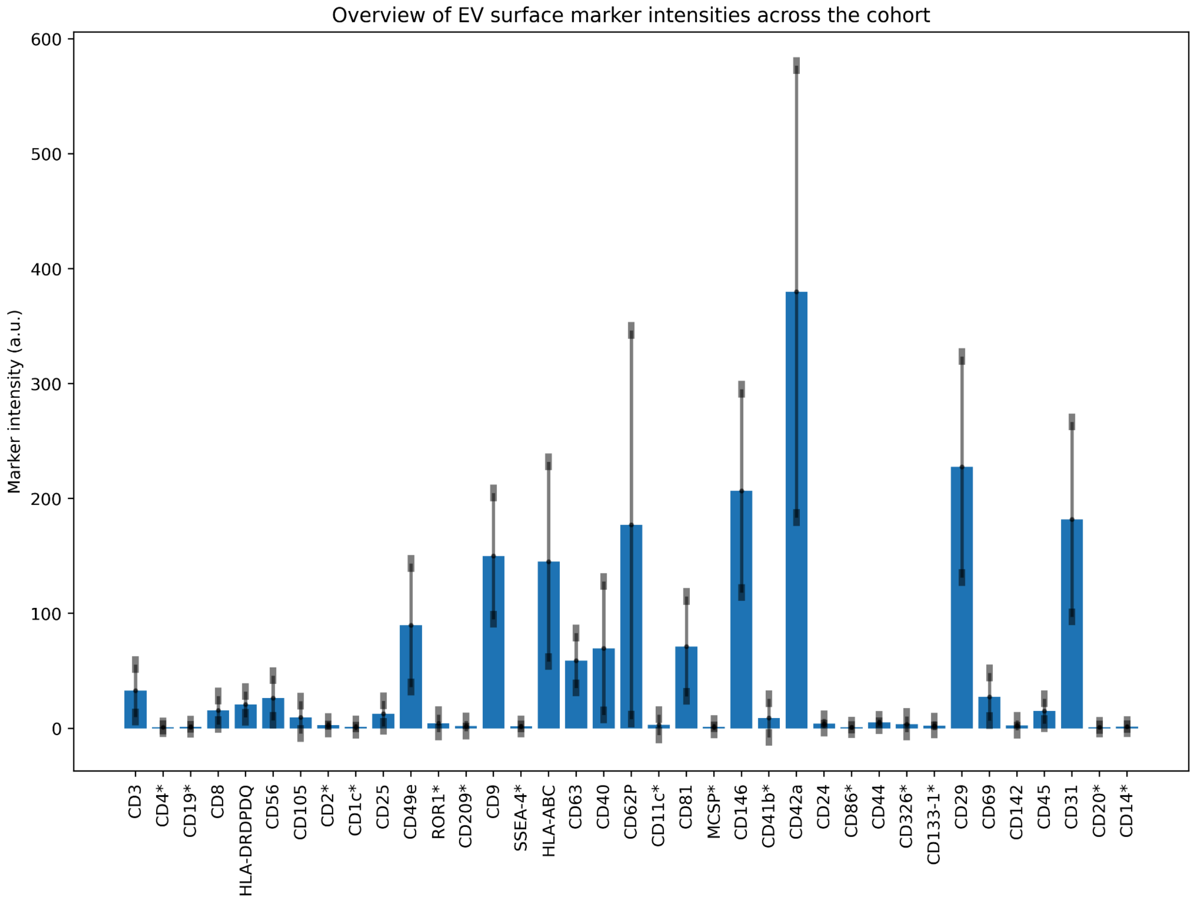


**Figure S1:** Mean fluorescence intensities of extracellular vesicle surface markers measured by flow cytometry, before log-transformation, averaged across all samples. Markers marked with an asterisk (*) yielded a detectable (non-zero) signal in fewer than 75% of samples and were therefore excluded from downstream analyses.

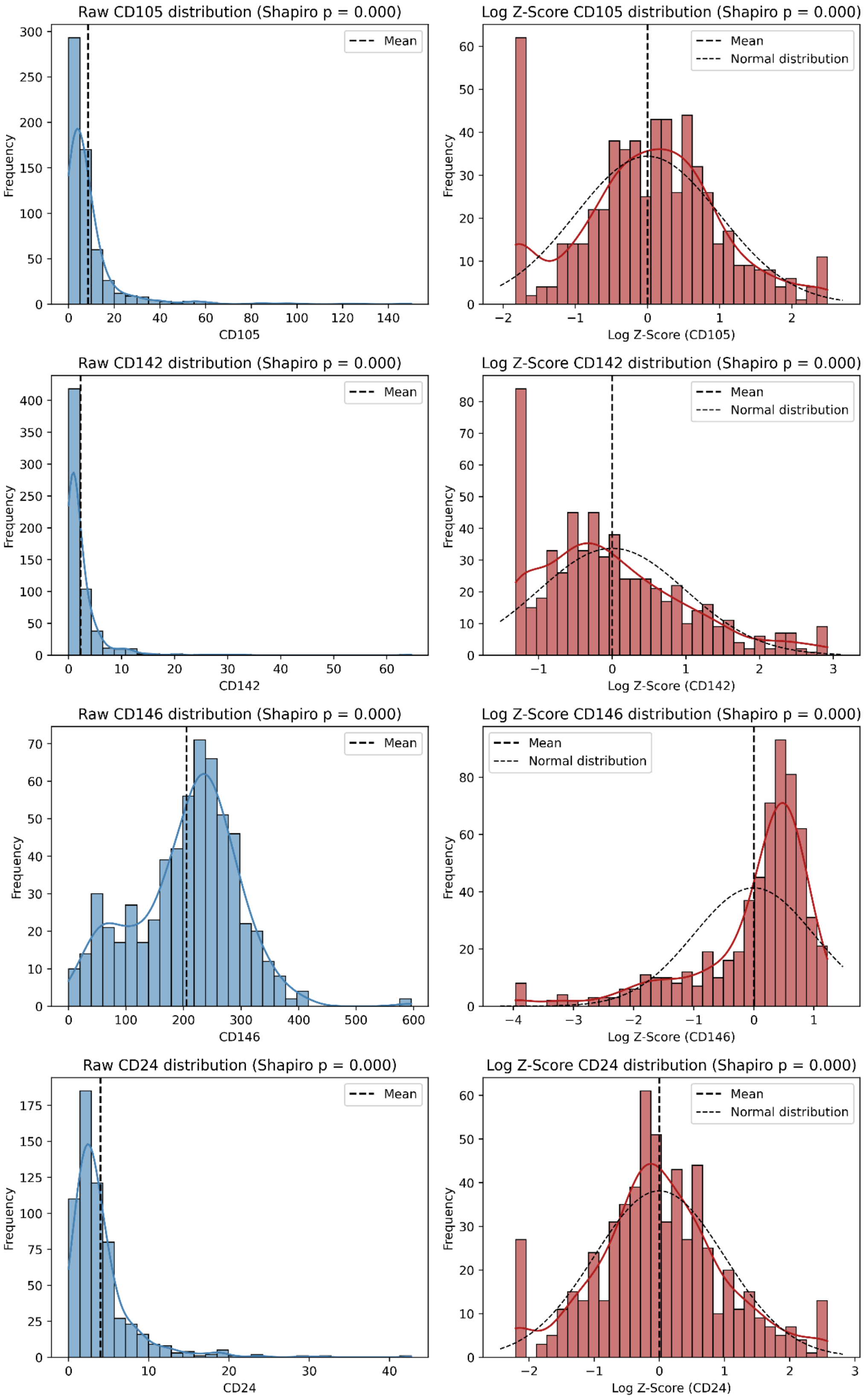


**Figure S2a**: Raw and log Z-score distributions of 22 EV surface marker fluorescence intensities. Each panel shows the raw measurement (left) alongside the log$_1$p-transformed, winsorised, and ComBat batch-corrected z-score (right). Shapiro-Wilk p-values are indicated in the panel titles. Showing markers CD105, CD142, CD146 and CD24.

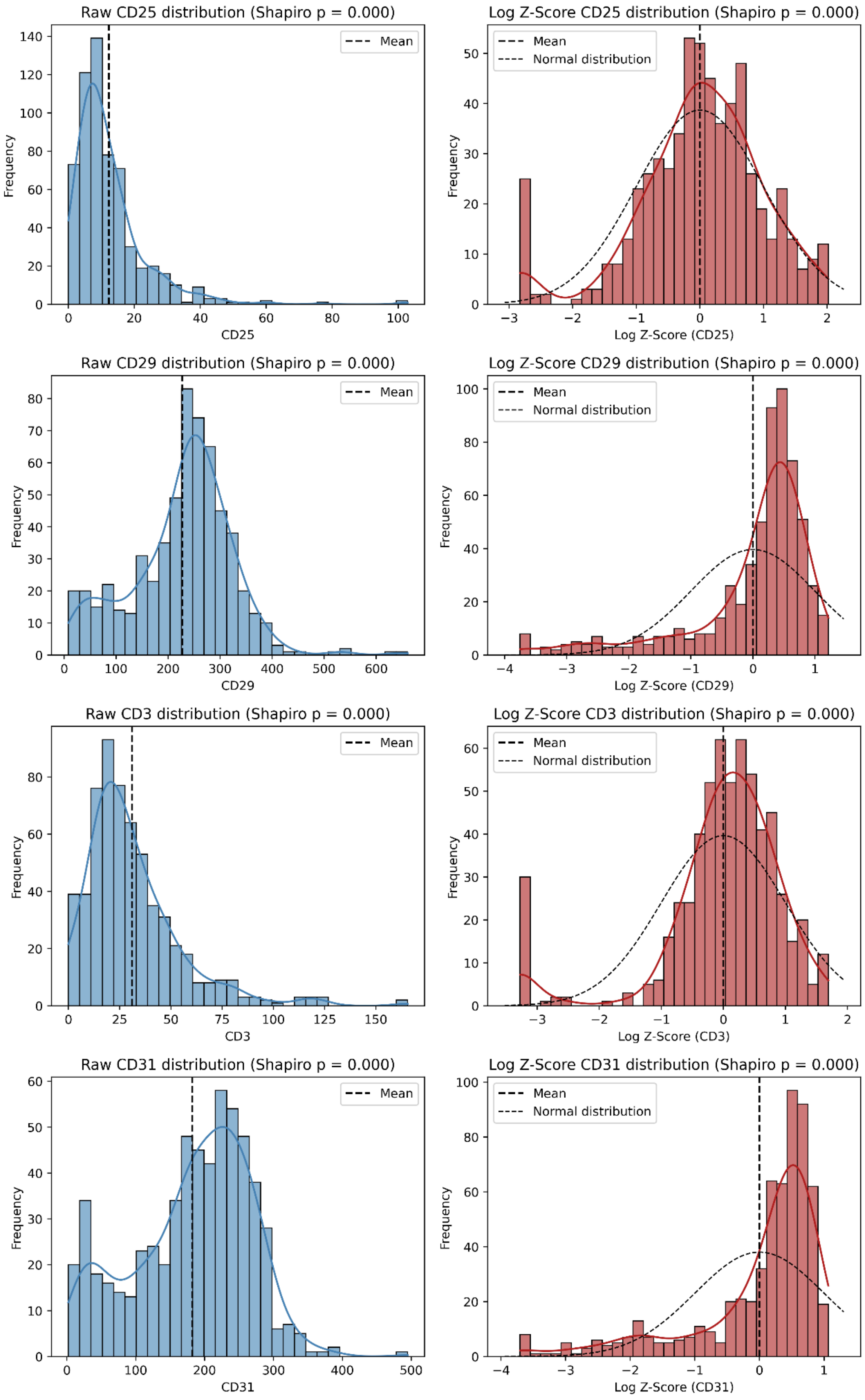


**Figure S2b**: As Figure S2a, showing markers CD25, CD29, CD3 and CD31.

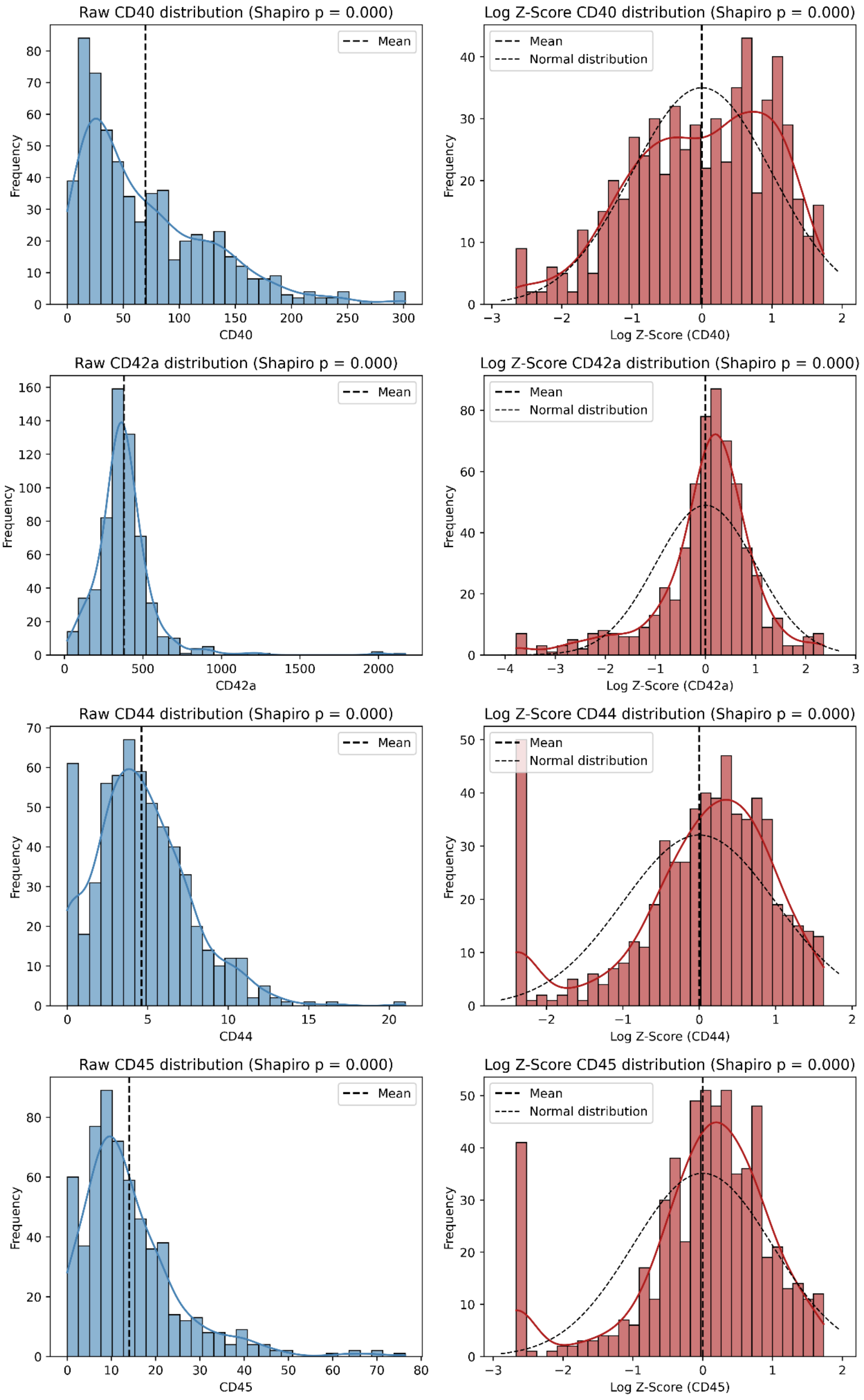


**Figure S2c**: As Figure S2a, showing markers CD40, CD42a, CD44 and CD45.

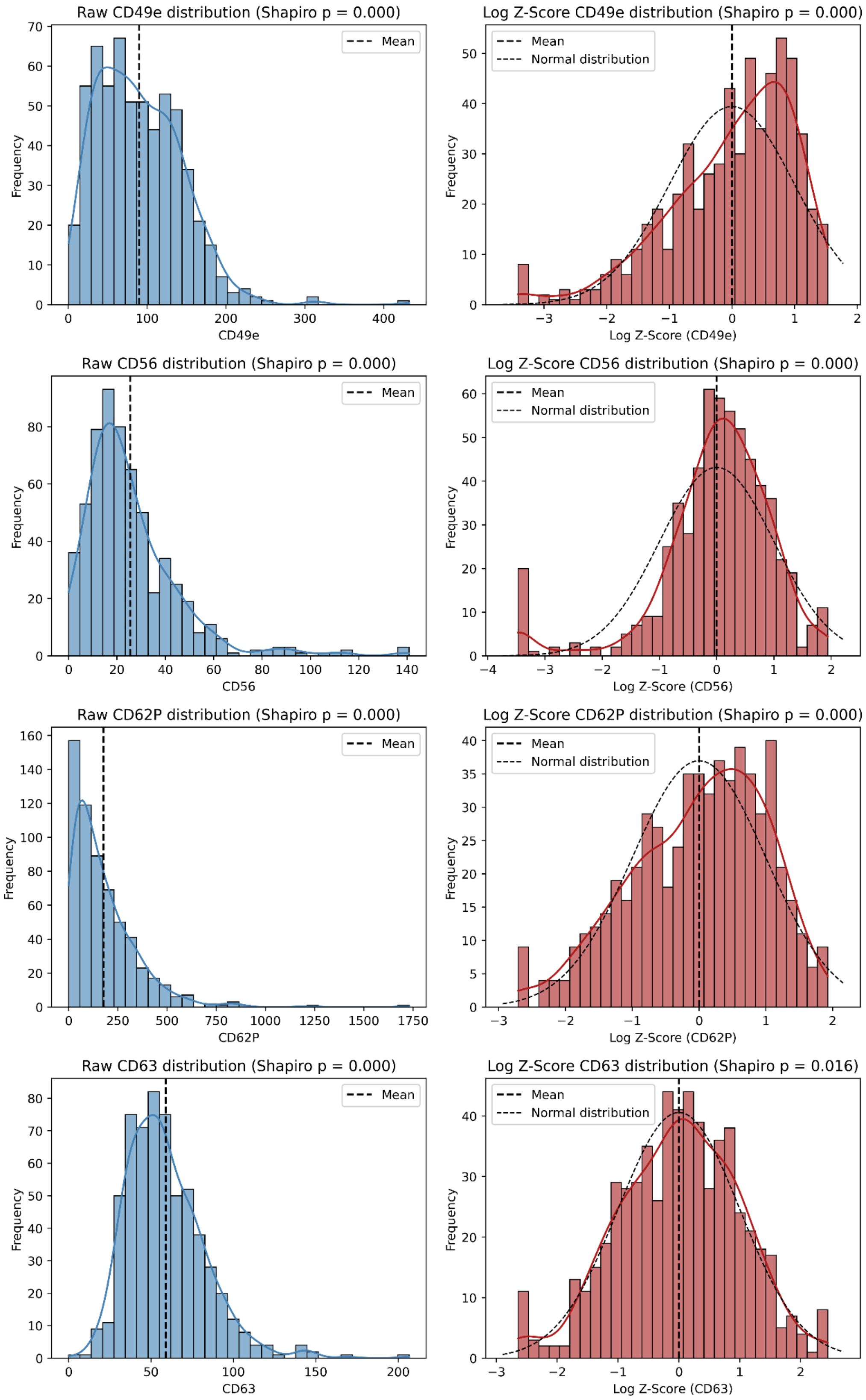


**Figure S2d**: As Figure S2a, showing markers CD49e, CD56, CD62P and CD63.

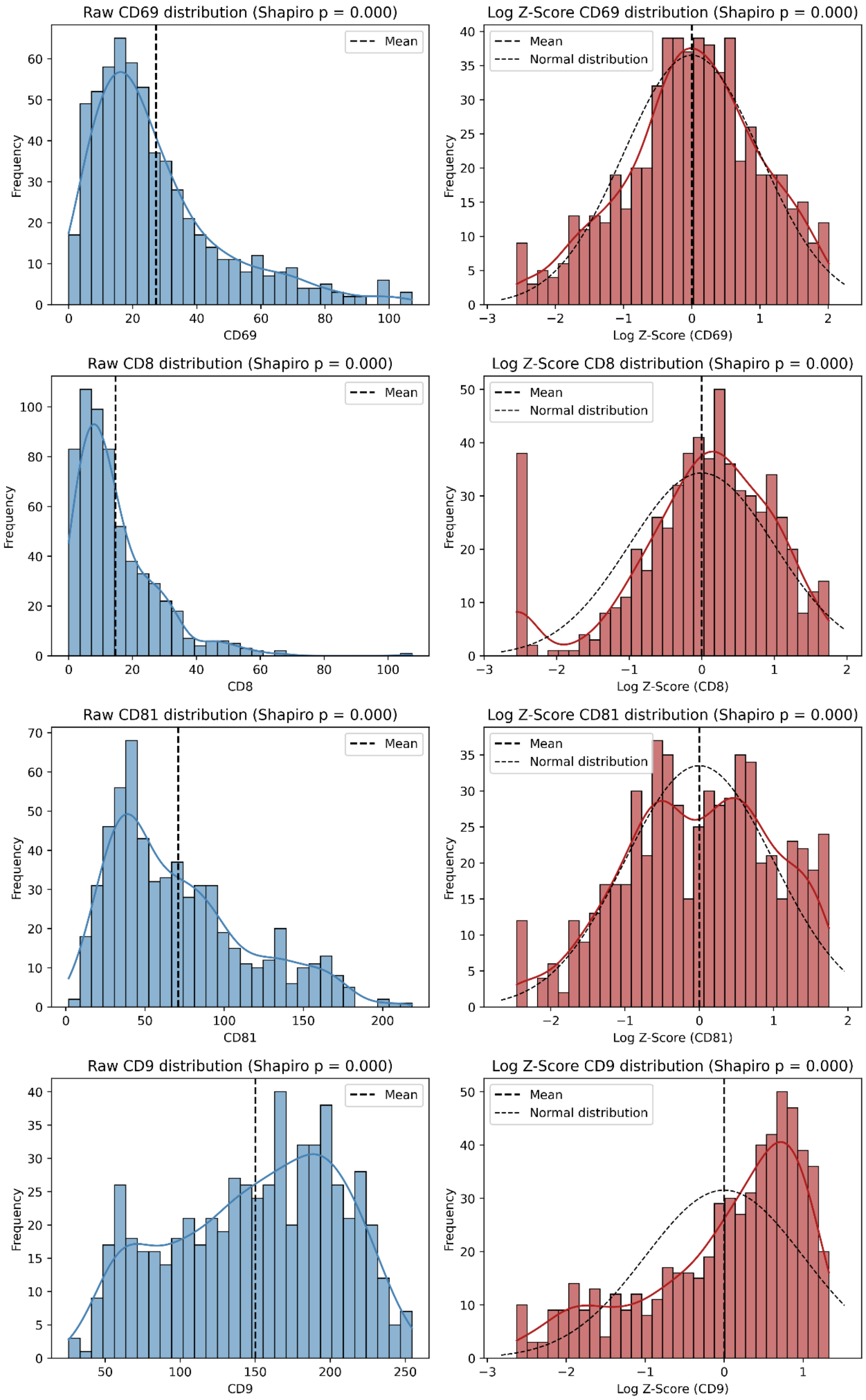


**Figure S2e**: As Figure S2a, showing markers CD69, CD8, CD81 and CD9.

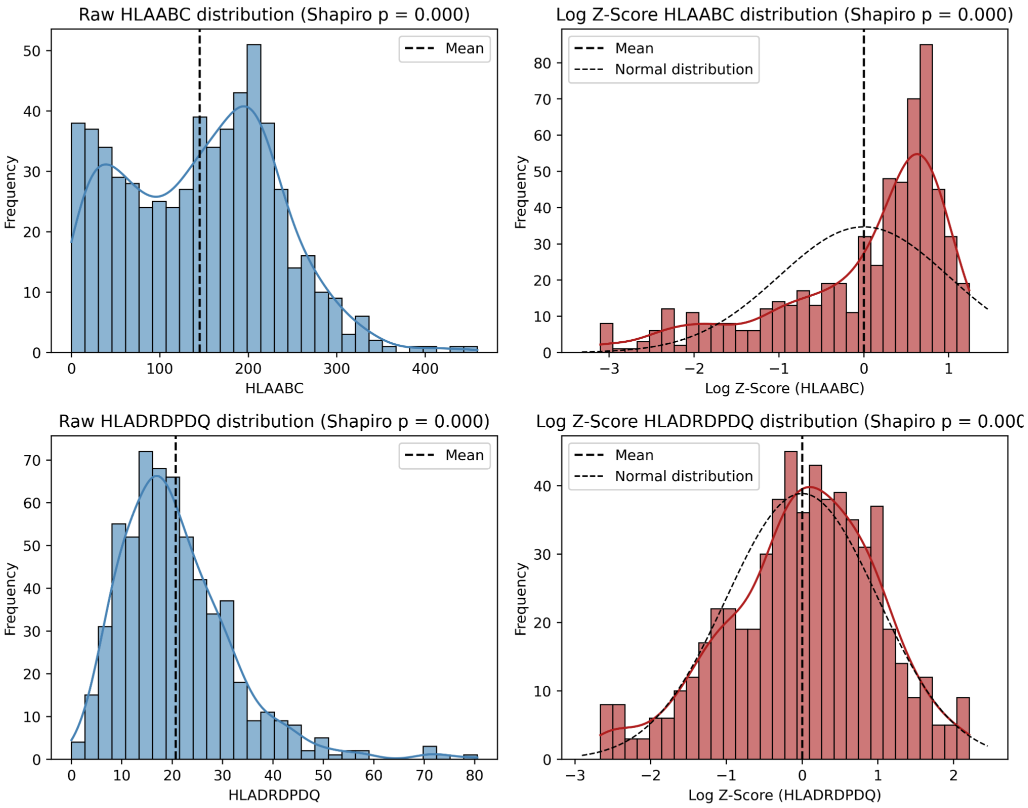


**Figure S2f**: As Figure S2a, showing markers HLA-ABC and HLA-DR/DP/DQ.

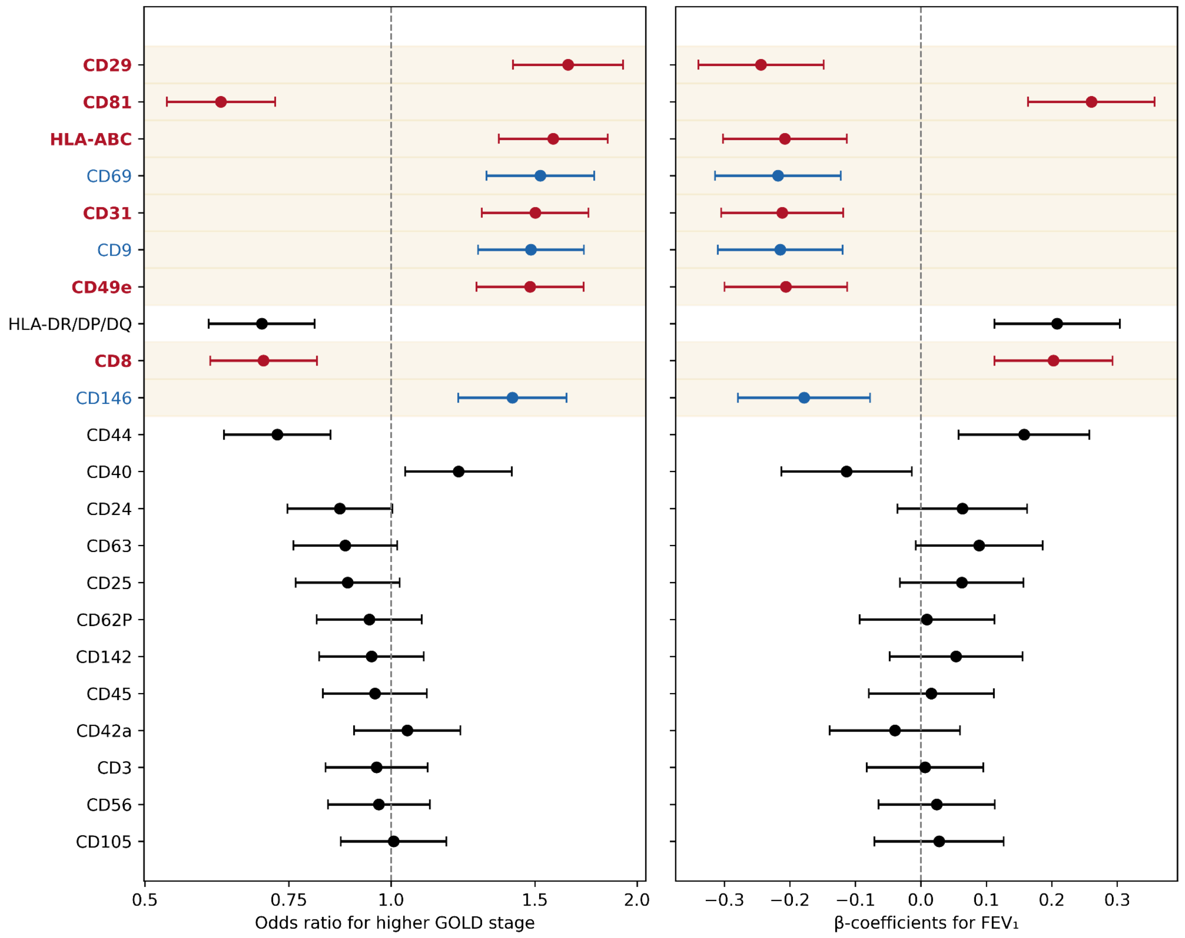


**Figure S3a. Cross-sectional sensitivity analysis without covariates.** Forest plots paralleling Figure 2: odds ratios for higher GOLD stage (left, ordinal logistic regression) and β-coefficients for $FEV_1$ z-score (right, linear regression) with 95% CI for all 22 markers in the full GOLD 0–4 cohort, fitted without covariate adjustment. Red: top-tier markers from the primary analysis (CD8, CD29, CD31, CD49e, CD81, HLA-ABC; see Figure 2). Significance reported as raw p-values (Table S3b.2).

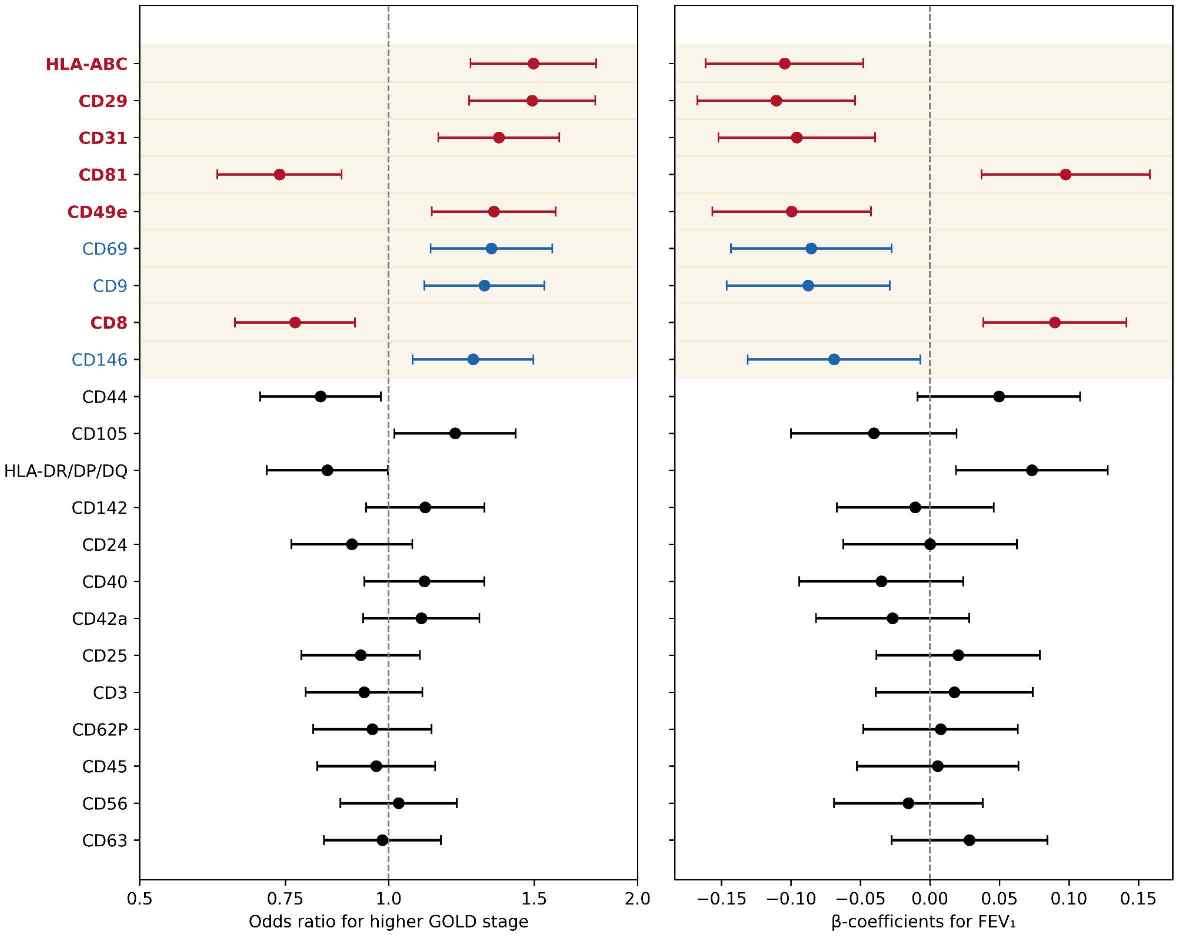


**Figure S3b. Cross-sectional sensitivity analysis with additional adjustment for FVC z-score.** Forest plots paralleling Figure 2: odds ratios for higher GOLD stage (left, ordinal logistic regression) and β-coefficients for $FEV_1$ z-score (right, linear regression) with 95% CI for all 22 markers in the full GOLD 0–4 cohort, with FVC z-score added to the prespecified covariate set. Red: top-tier markers from the primary analysis (CD8, CD29, CD31, CD49e, CD81, HLA-ABC; see Figure 2). Significance reported as raw p-values (Table S3b.3).

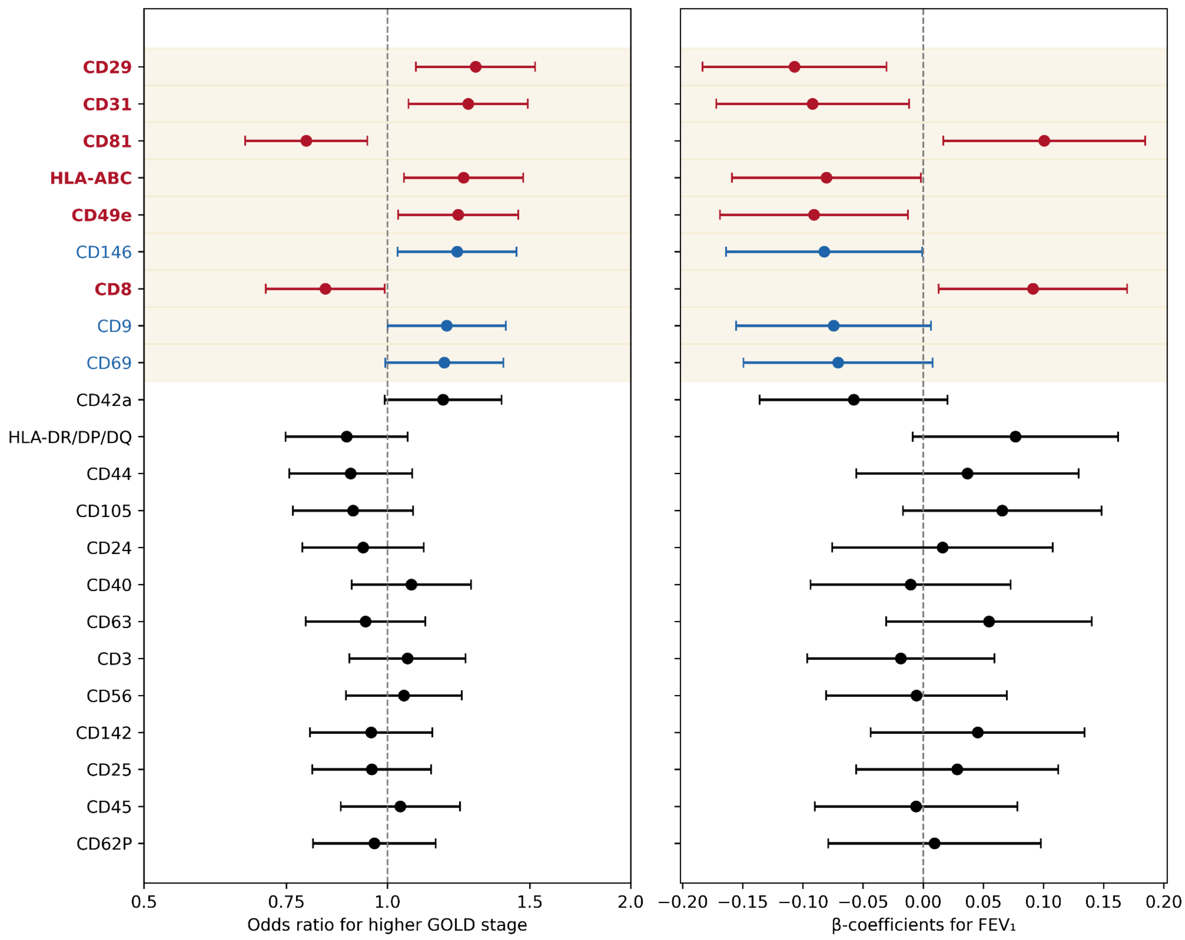


**Figure S3c. Cross-sectional sensitivity analysis restricted to established COPD (GOLD 1–4).** Forest plots paralleling Figure 2: odds ratios for higher GOLD stage (left, ordinal logistic regression) and β-coefficients for $FEV_1$ z-score (right, linear regression) with 95% CI for all 22 markers in the GOLD 1–4 subgroup, with the prespecified covariate set. Red: top-tier markers from the primary analysis (CD8, CD29, CD31, CD49e, CD81, HLA-ABC; see Figure 2). Significance reported as raw p-values (Table S3b.4).

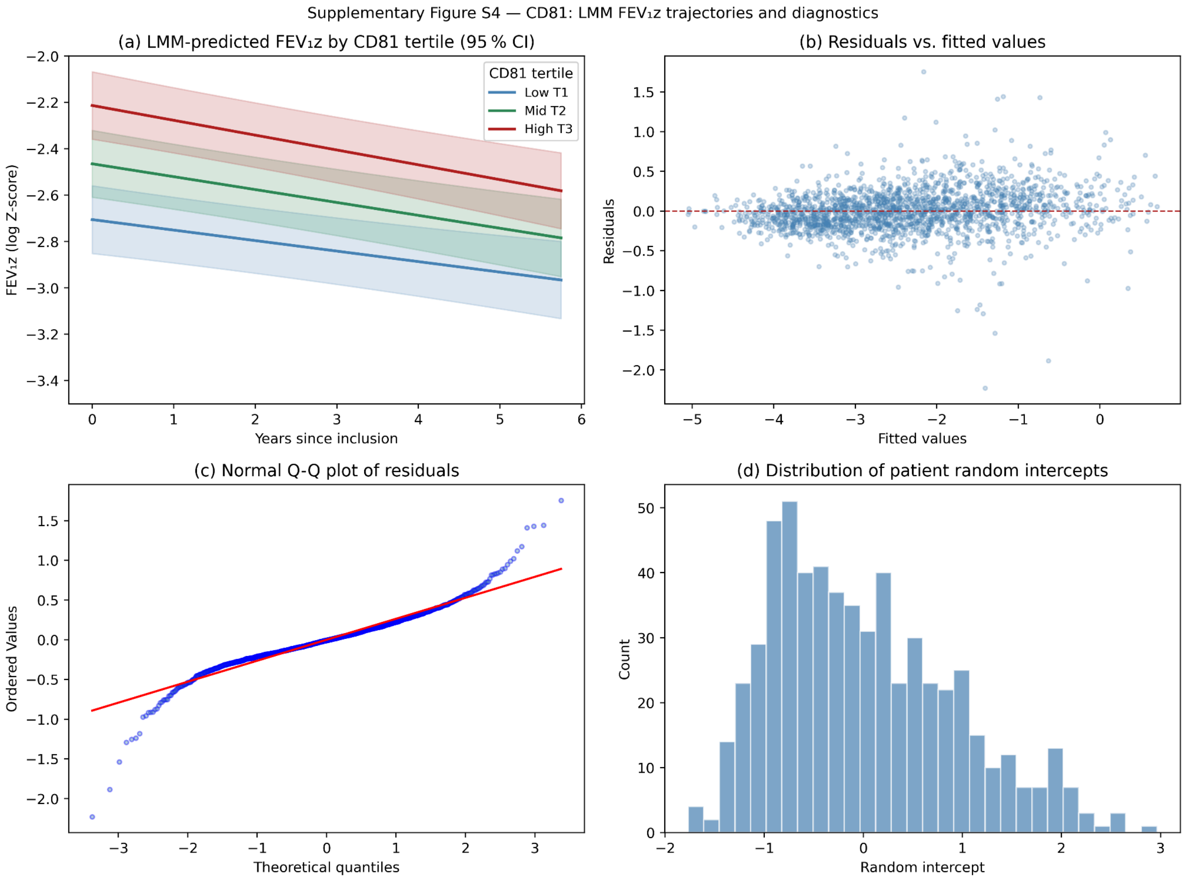


**Figure S4:** LMM-based $FEV_1z$ diagnostics for CD81. (a) LMM-predicted $FEV_1z$ trajectories by by baseline CD81 tertile (T1 = lowest CD81, T3 = highest CD81) plotted over years since inclusion; shaded bands show 95 % confidence intervals from the fixed-effects covariance matrix, with continuous covariates held at their median and binary covariates at their mode. (b) Residuals vs. fitted values. (c) Normal Q-Q plot of model residuals. (d) Distribution of patient-level random intercepts. Model: $FEV_1z \sim$ years × CD81 tertile + sex + age + BMI + smoking + pack-years + years with COPD + SGRQ activity + (1 + years | patient), fitted by REML.

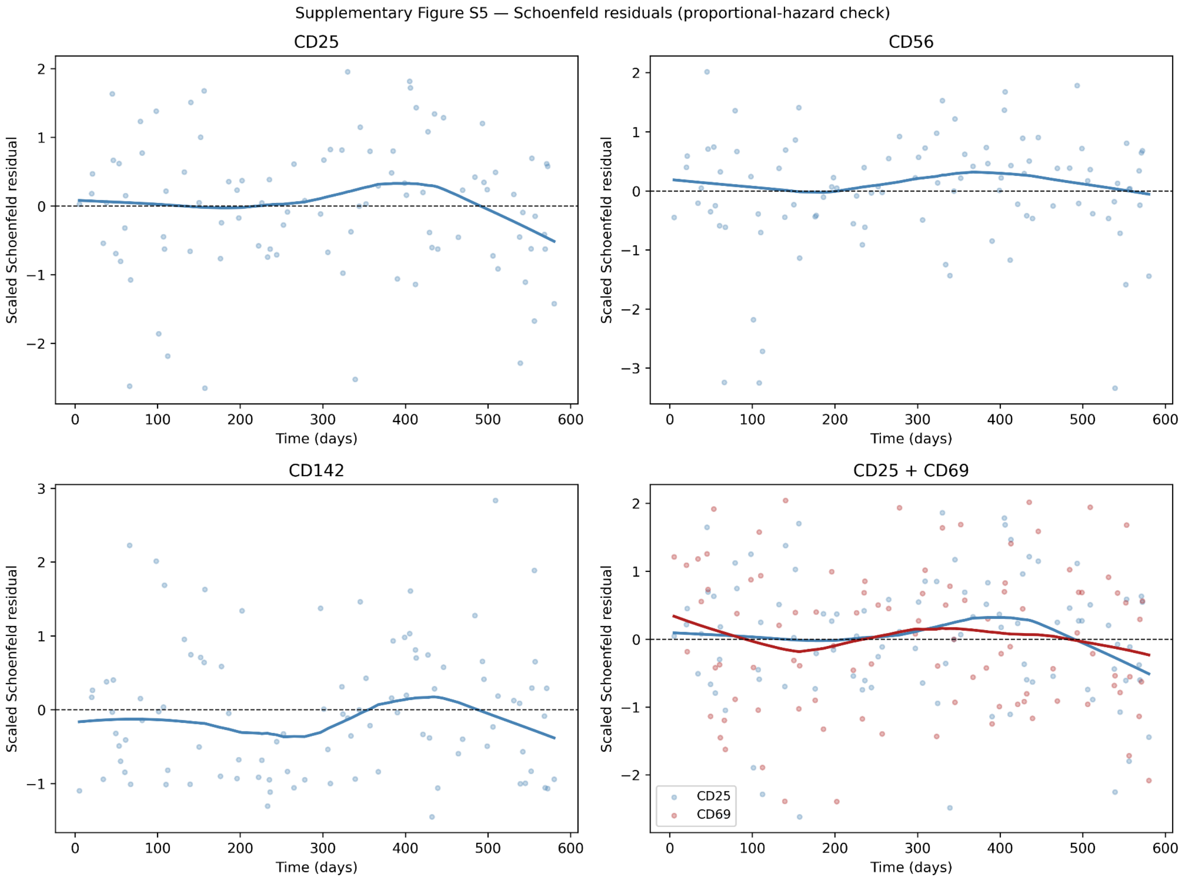


**Figure S5:** Scaled Schoenfeld residuals for the four primary Cox models (clinical core + $FEV_1z$ + EV marker) plotted against follow-up time. The LOWESS-smoothed trend (solid line) should be approximately flat if the proportional-hazards assumption holds; a systematic slope indicates a time-varying effect.

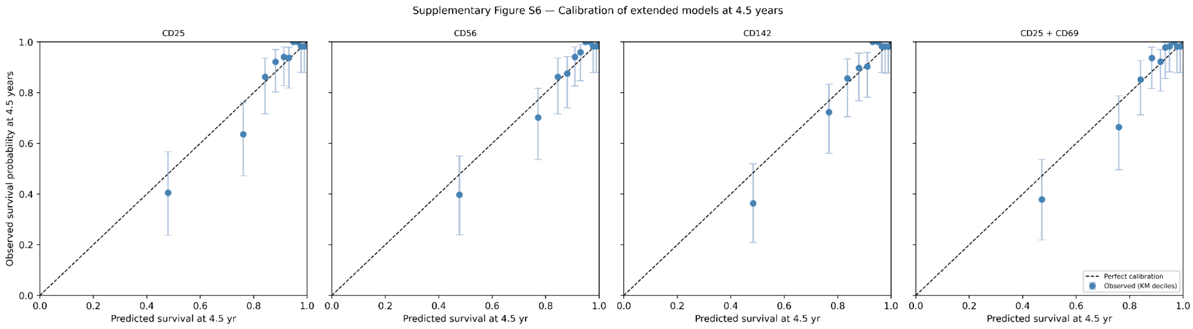


**Figure S6:** Calibration curves at 54 months for each extended model (clinical core + $FEV_1z$ + EV marker). Dots represent deciles of predicted survival probability; the diagonal indicates perfect calibration. Error bars show 95% confidence intervals from the Kaplan-Meier estimator.

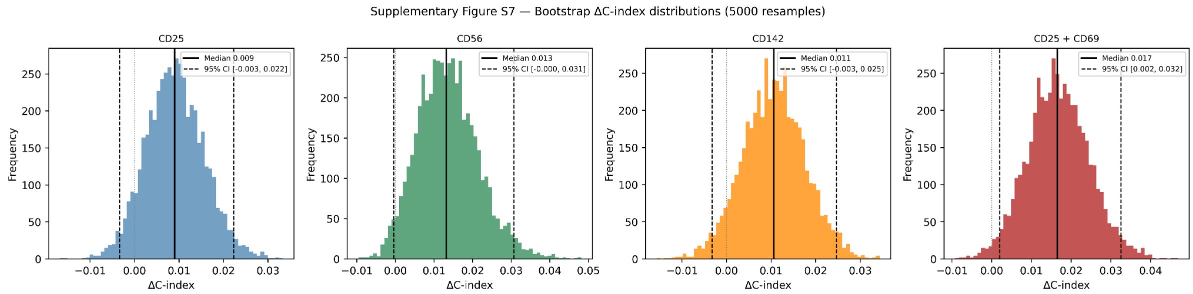


**Figure S7:** Bootstrap distributions of ΔC-index (extended minus clinical-only model) from 5000 resamples for each of the four primary EV-marker configurations. Solid vertical line: observed median ΔC; dashed lines: 2.5th and 97.5th percentile (95% bootstrap CI). The dotted line at 0 indicates no improvement in discrimination.

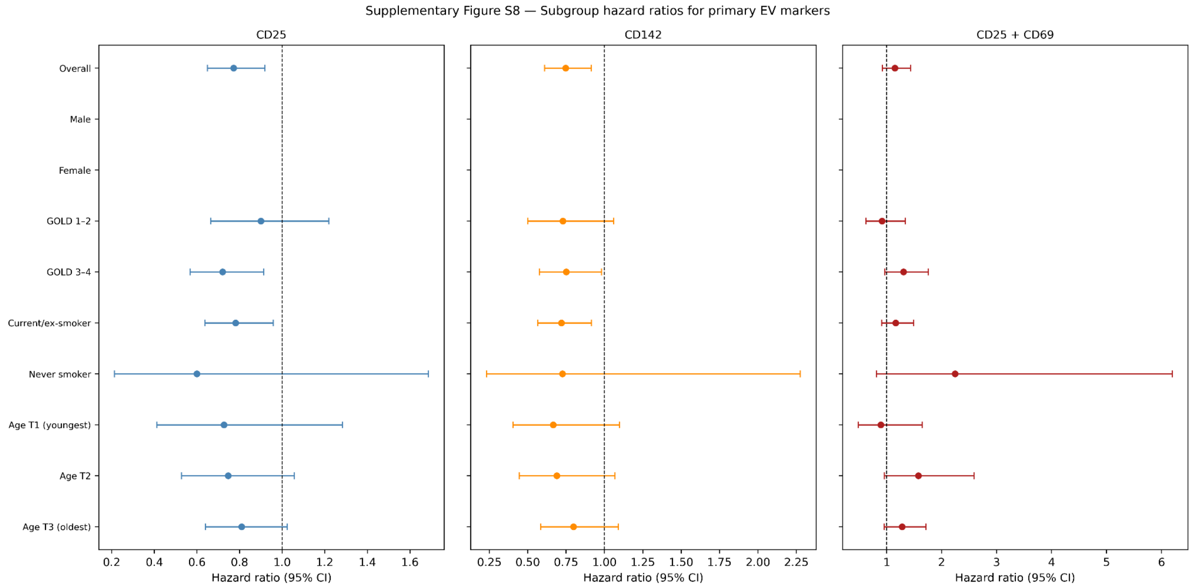


**Figure S8.** Subgroup hazard ratios for primary EV markers. Adjusted hazard ratios (95% CI) for CD25, CD142, and the CD25 + CD69 combination across sex, spirometric severity (GOLD 1-2 vs 3-4, or $FEV_1z$ tertile when GOLD was unavailable), smoking status (current/ex- vs never-smoker), and age tertile. All models adjusted for the Core covariate set (sex, age, leukocytes, $PaCO_2$, platelets, $FEV_1z$). Dashed line: HR = 1. Subgroup sample sizes were small; analyses are exploratory checks of homogeneity, not formal interaction tests.

### Tables

**Table S1.** Covariates and sample sizes per analysis. Core covariates entered into each model, with the corresponding number of patients, events, and (for the longitudinal model) repeated $FEV_1$ measurements. $FEV_1$ z-score was added to the core set in the primary cross-sectional and survival models. IL-6 was included in the cross-sectional ordinal and linear models but not in the linear mixed-effects model, due to aborted model convergence; the mixed model was therefore fitted on 597 patients contributing 1884 longitudinal $FEV_1$ measurements. The Cox model was fitted on 567 complete cases (98 events). Abbreviations: BMI, body mass index; SGRQ, St George's Respiratory Questionnaire; $PaCO_2$, arterial $CO_2$ partial pressure.

| | Covariates | Patients | Events | Measurements |
|---|---|---|---|---|
| Ordinal regression | Sex, age, BMI, smoking state, pack-years, years COPD, SGRQ activity, IL6 | 600 | - | - |
| Linear regression | | 600 | - | - |
| Linear mixed-effects model regression | Sex, age, BMI, smoking state, pack-years, years COPD, SGRQ activity | 597 | - | 1884 |
| Cox regression | sex, age, leukocytes, $PaCO_2$, platelets | 567 | 98 | - |

**Table S2: EV surface markers excluded from analysis due to low detectability.** The 15 markers listed yielded a detectable (non-zero) signal in fewer than 75% of cohort samples (n = 600) and were therefore excluded from all downstream analyses, leaving 22 markers for analysis. Detectable, n (%): number and percentage of samples in which the marker yielded a non-zero signal. Mean fluorescence intensities for all profiled markers, with the excluded markers indicated by an asterisk, are shown in Figure S1.

| **Marker** | **Prevalence** |
|---|---|
| CD4 | 169 (28.17) |
| CD19 | 251 (41.83) |
| CD2 | 439 (73.17) |
| CD1c | 175 (29.17) |
| ROR1 | 374 (62.33) |
| CD209 | 306 (51.00) |
| SSEA-4 | 106 (17.67) |
| CD11c | 395 (65.83) |
| MCSP | 143 (23.83) |
| CD41b | 97 (16.17) |
| CD86 | 182 (30.33) |
| CD326 | 380 (63.33) |
| CD133-1 | 332 (55.33) |
| CD20 | 207 (34.50) |
| CD14 | 294 (49.00) |

**Table S3a.** Cross-sectional EV marker associations across sensitivity analyses. **OR**: odds ratio for higher GOLD stage (ordinal logistic regression). **β**: coefficient for $FEV_1$ z-score (linear regression). Both per standard-deviation increase in marker level. **q**: Benjamini-Hochberg-adjusted p. **p**: raw p-values for sensitivity analysis. **Bold**: top-tier markers. Full per-scenario results in Table S3b. GOLD 1–4 without covariates is shown only in Table S3b.5 for space

| **Marker** | **Full range** (GOLD 0–4) | | | | | | **GOLD 1–4** | | | |
|---|---|---|---|---|---|---|---|---|---|---|
| | **Covariates (primary)** | | **No covariates** | | **+FVC** | | **Covariates** | | **+RV/TLC** | |
| | **OR (q)** | **β (q)** | **OR (p)** | **β (p)** | **OR (p)** | **β (p)** | **OR (p)** | **β (p)** | **OR (p)** | **β (p)** |
| CD105 | 1.0471 (0.7014) | 0.0289 (0.6691) | 1.0075 (0.9219) | 0.0279 (0.5793) | 1.2031 (0.0316) | -0.0403 (0.1838) | 0.9068 (0.2624) | 0.0657 (0.1184) | 0.9304 (0.4254) | 0.0574 (0.1920) |
| CD142 | 1.0049 (0.9566) | 0.0336 (0.6478) | 0.9467 (0.4664) | 0.0539 (0.2973) | 1.1073 (0.2251) | -0.0106 (0.7134) | 0.9547 (0.6016) | 0.0453 (0.3187) | 0.9468 (0.5454) | 0.0487 (0.2892) |
| CD146 | 1.3463 (0.0009) | -0.1383 (0.0029) | 1.4075 (<0.0001) | -0.1782 (0.0005) | 1.2653 (0.0060) | -0.0690 (0.0296) | 1.2194 (0.0221) | -0.0822 (0.0480) | 1.2005 (0.0386) | -0.0772 (0.0698) |
| CD24 | 0.8943 (0.3239) | 0.0116 (0.8745) | 0.8659 (0.0559) | 0.0635 (0.2084) | 0.9028 (0.2341) | 0.0000 (0.9988) | 0.9330 (0.4316) | 0.0160 (0.7316) | 0.9265 (0.3983) | 0.0142 (0.7662) |
| CD25 | 0.9078 (0.3383) | 0.0258 (0.6944) | 0.8852 (0.1029) | 0.0626 (0.1930) | 0.9250 (0.3558) | 0.0203 (0.4989) | 0.9563 (0.6044) | 0.0283 (0.5095) | 0.9402 (0.4860) | 0.0339 (0.4469) |
| **CD29** | 1.5261 (<0.0001) | -0.1677 (0.0004) | 1.6463 (<0.0001) | -0.2440 (<0.0001) | 1.4914 (<0.0001) | -0.1105 (0.0001) | 1.2854 (0.0037) | -0.1070 (0.0061) | 1.2907 (0.0037) | -0.1079 (0.0058) |
| CD3 | 0.9693 (0.7982) | -0.0100 (0.8745) | 0.9603 (0.5797) | 0.0067 (0.8833) | 0.9339 (0.4089) | 0.0175 (0.5436) | 1.0588 (0.4986) | -0.0186 (0.6391) | 1.0499 (0.5747) | -0.0183 (0.6576) |
| **CD31** | 1.3804 (0.0005) | -0.1410 (0.0020) | 1.5013 (<0.0001) | -0.2115 (<0.0001) | 1.3591 (0.0004) | -0.0958 (0.0008) | 1.2587 (0.0079) | -0.0919 (0.0246) | 1.2553 (0.0097) | -0.0916 (0.0250) |

| **Marker** | **Full range** (GOLD 0–4) | | | | | | **GOLD 1–4** | | | |
|---|---|---|---|---|---|---|---|---|---|---|
| | **Covariates (primary)** | | **No covariates** | | **+FVC** | | **Covariates** | | **+RV/TLC** | |
| | **OR (q)** | **β (q)** | **OR (p)** | **β (p)** | **OR (p)** | **β (p)** | **OR (p)** | **β (p)** | **OR (p)** | **β (p)** |
| CD40 | 1.1039 (0.3383) | -0.0505 (0.3679) | 1.2093 (0.0131) | -0.1133 (0.0256) | 1.1045 (0.2430) | -0.0349 (0.2461) | 1.0705 (0.4317) | -0.0104 (0.8064) | 1.0466 (0.6054) | -0.0007 (0.9864) |
| CD42a | 1.1077 (0.3310) | -0.0636 (0.2189) | 1.0469 (0.5482) | -0.0395 (0.4372) | 1.0952 (0.2708) | -0.0268 (0.3400) | 1.1717 (0.0620) | -0.0578 (0.1470) | 1.1766 (0.0604) | -0.0627 (0.1204) |
| CD44 | 0.8201 (0.0347) | 0.0651 (0.2615) | 0.7261 (<0.0001) | 0.1577 (0.0020) | 0.8272 (0.0267) | 0.0496 (0.0960) | 0.9007 (0.2408) | 0.0368 (0.4341) | 0.8902 (0.2052) | 0.0384 (0.4302) |
| CD45 | 0.9957 (0.9566) | -0.0118 (0.8745) | 0.9557 (0.5428) | 0.0162 (0.7398) | 0.9660 (0.6794) | 0.0056 (0.8514) | 1.0376 (0.6695) | -0.0059 (0.8901) | 1.0230 (0.7994) | -0.0047 (0.9173) |
| **CD49e** | 1.3523 (0.0009) | -0.1468 (0.0014) | 1.4797 (<0.0001) | -0.2060 (<0.0001) | 1.3400 (0.0009) | -0.0994 (0.0006) | 1.2231 (0.0211) | -0.0908 (0.0229) | 1.2173 (0.0275) | -0.0900 (0.0275) |
| CD56 | 0.9917 (0.9566) | -0.0054 (0.9263) | 0.9664 (0.6412) | 0.0244 (0.5906) | 1.0283 (0.7353) | -0.0155 (0.5692) | 1.0481 (0.5763) | -0.0056 (0.8830) | 1.0429 (0.6224) | -0.0070 (0.8550) |
| CD62P | 0.9512 (0.7014) | 0.0019 (0.9660) | 0.9407 (0.4187) | 0.0094 (0.8578) | 0.9559 (0.5913) | 0.0077 (0.7854) | 0.9634 (0.6750) | 0.0095 (0.8336) | 0.9594 (0.6532) | 0.0145 (0.7579) |
| CD63 | 0.9399 (0.6032) | 0.0641 (0.2349) | 0.8792 (0.0844) | 0.0892 (0.0706) | 0.9826 (0.8326) | 0.0284 (0.3196) | 0.9395 (0.4730) | 0.0547 (0.2100) | 0.9414 (0.4994) | 0.0602 (0.1788) |
| CD69 | 1.3637 (0.0008) | -0.1269 (0.0046) | 1.5228 (<0.0001) | -0.2181 (<0.0001) | 1.3317 (0.0009) | -0.0855 (0.0037) | 1.1758 (0.0592) | -0.0708 (0.0774) | 1.1544 (0.1013) | -0.0657 (0.1080) |
| **CD8** | 0.7470 (0.0010) | 0.1315 (0.0029) | 0.6984 (<0.0001) | 0.2027 (<0.0001) | 0.7706 (0.0022) | 0.0898 (0.0006) | 0.8378 (0.0408) | 0.0912 (0.0225) | 0.8298 (0.0391) | 0.0871 (0.0389) |

| **Marker** | **Full range** (GOLD 0–4) | | | | | | **GOLD 1–4** | | | |
|---|---|---|---|---|---|---|---|---|---|---|
| | **Covariates (primary)** | | **No covariates** | | **+FVC** | | **Covariates** | | **+RV/TLC** | |
| | **OR (q)** | **β (q)** | **OR (p)** | **β (p)** | **OR (p)** | **β (p)** | **OR (p)** | **β (p)** | **OR (p)** | **β (p)** |
| **CD81** | 0.6958 (0.0001) | 0.1623 (0.0014) | 0.6195 (<0.0001) | 0.2605 (<0.0001) | 0.7381 (0.0006) | 0.0977 (0.0016) | 0.7937 (0.0093) | 0.1006 (0.0187) | 0.7941 (0.0110) | 0.1037 (0.0159) |
| CD9 | 1.3481 (0.0009) | -0.1353 (0.0029) | 1.4831 (<0.0001) | -0.2146 (<0.0001) | 1.3057 (0.0017) | -0.0876 (0.0034) | 1.1840 (0.0491) | -0.0744 (0.0720) | 1.1678 (0.0767) | -0.0695 (0.1009) |
| HLA-ABC | 1.4971 (<0.0001) | -0.1469 (0.0014) | 1.5791 (<0.0001) | -0.2075 (<0.0001) | 1.4963 (<0.0001) | -0.1045 (0.0003) | 1.242 (0.0125) | -0.0804 (0.0448) | 1.2417 (0.0146) | -0.0836 (0.0400) |
| HLA-DR/ DP/DQ | 0.8202 (0.0347) | 0.0971 (0.0503) | 0.6950 (<0.0001) | 0.2082 (<0.0001) | 0.8430 (0.0470) | 0.0734 (0.0085) | 0.891 (0.1916) | 0.0767 (0.0778) | 0.8930 (0.2163) | 0.0656 (0.1445) |

**Table S3b.1:** Cross-sectional associations of EV markers with COPD severity: primary analysis (full GOLD 0–4 cohort, prespecified covariates). OR: odds ratio for higher GOLD stage per standard-deviation marker increase (ordinal logistic regression). β: coefficient for $FEV_1$ z-score per SD (linear regression, HC3-robust SE). p / q: raw / Benjamini-Hochberg-adjusted (across 22 markers) p-values. **$R^2$**: McFadden's pseudo-$R^2$ for the ordinal model (left block) and the linear regression's $R^2$ for the right block. **$\Delta R^2$**: incremental marker contribution. **$pR^2$**: marker contribution to the linear regression. **Bold**: top-tier markers (CD8, CD29, CD31, CD49e, CD81, HLA-ABC; see Table S3a). Sensitivity analyses with raw p-values: Tables S3b.2-S3b.6.

| **Marker** | **Ordinal logistic regression** (GOLD stages) | | | | | | **Linear regression** ($FEV_1$ z-score) | | | |
|---|---|---|---|---|---|---|---|---|---|---|
| | **OR** (CI 95%) | **p** | **q** | **$R^2$** | **$\Delta R^2$** | **LR p** | **β** (95% CI) | **p** | **q** | **$pR^2$** |
| CD105 | 1.0471 (0.8918 to 1.2295) | 0.5739 | 0.7014 | 0.1438 | 0.0002 | 0.5741 | 0.0289 (-0.0525 to 0.1102) | 0.4866 | 0.6691 | 0.0006 |
| CD142 | 1.0049 (0.8565 to 1.1790) | 0.9524 | 0.9566 | 0.1436 | 0.0000 | 0.9530 | 0.0336 (-0.0520 to 0.1192) | 0.4417 | 0.6478 | 0.0008 |
| CD146 | 1.3463 (1.1442 to 1.5840) | 0.0003 | 0.0009 | 0.1518 | 0.0082 | 0.0003 | -0.1383 (-0.2211 to -0.0555) | 0.0011 | 0.0029 | 0.0129 |
| CD24 | 0.8943 (0.7605 to 1.0516) | 0.1767 | 0.3239 | 0.1447 | 0.0011 | 0.1763 | 0.0116 (-0.0758 to 0.0990) | 0.7950 | 0.8745 | 0.0001 |
| CD25 | 0.9078 (0.7750 to 1.0633) | 0.2306 | 0.3383 | 0.1445 | 0.0009 | 0.2307 | 0.0258 (-0.0560 to 0.1076) | 0.5366 | 0.6944 | 0.0004 |
| **CD29** | 1.5261 (1.2936 to 1.8005) | <0.0001 | <0.0001 | 0.1597 | 0.0161 | <0.0001 | -0.1677 (-0.2440 to -0.0915) | <0.0001 | 0.0004 | 0.0191 |
| CD3 | 0.9693 (0.8320 to 1.1293) | 0.6894 | 0.7982 | 0.1437 | 0.0001 | 0.6892 | -0.0100 (-0.0851 to 0.0651) | 0.7940 | 0.8745 | 0.0001 |
| **CD31** | 1.3804 (1.1753 to 1.6213) | <0.0001 | 0.0005 | 0.1534 | 0.0098 | <0.0001 | -0.1410 (-0.2198 to -0.0622) | 0.0005 | 0.0020 | 0.0134 |
| CD40 | 1.1039 (0.9400 to 1.2963) | 0.2281 | 0.3383 | 0.1445 | 0.0009 | 0.2278 | -0.0505 (-0.1336 to 0.0327) | 0.2341 | 0.3679 | 0.0017 |
| CD42a | 1.1077 (0.9488 to 1.2932) | 0.1956 | 0.3310 | 0.1446 | 0.0011 | 0.1960 | -0.0636 (-0.1414 to 0.0143) | 0.1094 | 0.2189 | 0.0028 |
| CD44 | 0.8201 (0.6965 to 0.9657) | 0.0174 | 0.0347 | 0.1472 | 0.0036 | 0.0172 | 0.0651 (-0.0245 to 0.1548) | 0.1545 | 0.2615 | 0.0028 |
| CD45 | 0.9957 (0.8510 to 1.1649) | 0.9566 | 0.9566 | 0.1436 | 0.0000 | 0.9570 | -0.0118 (-0.0933 to 0.0697) | 0.7768 | 0.8745 | 0.0001 |
| **CD49e** | 1.3523 (1.1485 to 1.5922) | 0.0003 | 0.0009 | 0.1519 | 0.0083 | 0.0003 | -0.1468 (-0.2244 to -0.0693) | 0.0002 | 0.0014 | 0.0141 |

| **Marker** | **Ordinal logistic regression** (GOLD stages) | | | | | | **Linear regression** ($FEV_1$ z-score) | | | |
|---|---|---|---|---|---|---|---|---|---|---|
| | **OR** (CI 95%) | **p** | **q** | **R²** | **ΔR²** | **LR p** | **β** (95% CI) | **p** | **q** | **pR²** |
| CD56 | 0.9917 (0.8518 to 1.1545) | 0.9144 | 0.9566 | 0.1436 | 0.0000 | 0.9143 | -0.0054 (-0.0783 to 0.0675) | 0.8842 | 0.9263 | 0.0000 |
| CD62P | 0.9512 (0.8098 to 1.1174) | 0.5428 | 0.7014 | 0.1438 | 0.0002 | 0.5427 | 0.0019 (-0.0843 to 0.0880) | 0.9660 | 0.9660 | 0.0000 |
| CD63 | 0.9399 (0.8034 to 1.0995) | 0.4387 | 0.6032 | 0.1440 | 0.0004 | 0.4387 | 0.0641 (-0.0185 to 0.1467) | 0.1281 | 0.2349 | 0.0028 |
| CD69 | 1.3637 (1.1592 to 1.6042) | 0.0002 | 0.0008 | 0.1525 | 0.0089 | 0.0002 | -0.1269 (-0.2069 to -0.0469) | 0.0019 | 0.0046 | 0.0109 |
| **CD8** | 0.7470 (0.6358 to 0.8776) | 0.0004 | 0.0010 | 0.1516 | 0.0080 | 0.0004 | 0.1315 (0.0535 to 0.2094) | 0.0009 | 0.0029 | 0.0116 |
| **CD81** | 0.6958 (0.5897 to 0.8211) | <0.0001 | 0.0001 | 0.1554 | 0.0118 | <0.0001 | 0.1623 (0.0781 to 0.2466) | 0.0002 | 0.0014 | 0.0172 |
| CD9 | 1.3481 (1.1493 to 1.5814) | 0.0002 | 0.0009 | 0.1521 | 0.0085 | 0.0002 | -0.1353 (-0.2162 to -0.0545) | 0.0010 | 0.0029 | 0.0123 |
| HLA-ABC | 1.4971 (1.2690 to 1.7662) | <0.0001 | <0.0001 | 0.1582 | 0.0146 | <0.0001 | -0.1469 (-0.2255 to -0.0683) | 0.0002 | 0.0014 | 0.0143 |
| HLA-DR/ DP/DQ | 0.8202 (0.6967 to 0.9657) | 0.0173 | 0.0347 | 0.1472 | 0.0036 | 0.0170 | 0.0971 (0.0135 to 0.1806) | 0.0229 | 0.0503 | 0.0060 |

**Table S3b.2: Cross-sectional associations of EV markers with COPD severity: sensitivity analysis without covariates (full GOLD 0–4 cohort). OR**: odds ratio for higher GOLD stage per standard-deviation marker increase (ordinal logistic regression). **β**: coefficient for $FEV_1$ z-score per SD (linear regression, HC3-robust SE). **p**: raw p-value. **R²**: McFadden's pseudo-$R^2$ for the ordinal model. Significance is reported as raw p-values; FDR adjustment is applied only in the primary analysis (Table S3b.1). **Bold**: top-tier markers (CD8, CD29, CD31, CD49e, CD81, HLA-ABC; see Table S3a).

| **Marker** | **Ordinal logistic regression** (GOLD stages) | | | **Linear regression** ($FEV_1$ z-score) | |
|---|---|---|---|---|---|
| | **OR** (CI 95%) | **p** | **R²** | **β** (95% CI) | **p** |
| CD105 | 1.0075 (0.8684 to 1.1688) | 0.9219 | 0.0000 | 0.0279 (-0.0708 to 0.1266) | 0.5793 |
| CD142 | 0.9467 (0.8169 to 1.0971) | 0.4664 | 0.0003 | 0.0539 (-0.0475 to 0.1554) | 0.2973 |
| CD146 | 1.4075 (1.2087 to 1.6390) | <0.0001 | 0.0113 | -0.1782 (-0.2790 to -0.0774) | 0.0005 |
| CD24 | 0.8659 (0.7470 to 1.0036) | 0.0559 | 0.0021 | 0.0635 (-0.0354 to 0.1625) | 0.2084 |
| CD25 | 0.8852 (0.7646 to 1.0249) | 0.1029 | 0.0016 | 0.0626 (-0.0316 to 0.1568) | 0.1930 |
| **CD29** | 1.6463 (1.4099 to 1.9225) | <0.0001 | 0.0235 | -0.2440 (-0.3396 to -0.1484) | <0.0001 |
| CD3 | 0.9603 (0.8319 to 1.1084) | 0.5797 | 0.0002 | 0.0067 (-0.0823 to 0.0956) | 0.8833 |
| **CD31** | 1.5013 (1.2918 to 1.7448) | <0.0001 | 0.0165 | -0.2115 (-0.3049 to -0.1182) | <0.0001 |
| CD40 | 1.2093 (1.0408 to 1.4051) | 0.0131 | 0.0036 | -0.1133 (-0.2128 to -0.0138) | 0.0256 |
| CD42a | 1.0469 (0.9014 to 1.2160) | 0.5482 | 0.0002 | -0.0395 (-0.1390 to 0.0601) | 0.4372 |
| CD44 | 0.7261 (0.6250 to 0.8436) | <0.0001 | 0.0104 | 0.1577 (0.0578 to 0.2576) | 0.0020 |
| CD45 | 0.9557 (0.8258 to 1.1059) | 0.5428 | 0.0002 | 0.0162 (-0.0793 to 0.1116) | 0.7398 |
| **CD49e** | 1.4797 (1.2723 to 1.7210) | <0.0001 | 0.0152 | -0.2060 (-0.2995 to -0.1124) | <0.0001 |
| CD56 | 0.9664 (0.8372 to 1.1156) | 0.6412 | 0.0001 | 0.0244 (-0.0644 to 0.1131) | 0.5906 |

| **Marker** | **Ordinal logistic regression** (GOLD stages) | | | **Linear regression** ($FEV_1$ z-score) | |
|---|---|---|---|---|---|
| | **OR** (CI 95%) | **p** | **$R^2$** | **β** (95% CI) | **p** |
| CD62P | 0.9407 (0.8112 to 1.0909) | 0.4187 | 0.0004 | 0.0094 (-0.0935 to 0.1123) | 0.8578 |
| CD63 | 0.8792 (0.7596 to 1.0176) | 0.0844 | 0.0017 | 0.0892 (-0.0075 to 0.1859) | 0.0706 |
| CD69 | 1.5228 (1.3083 to 1.7725) | <0.0001 | 0.0174 | -0.2181 (-0.3140 to -0.1222) | <0.0001 |
| **CD8** | 0.6984 (0.6010 to 0.8116) | <0.0001 | 0.0132 | 0.2027 (0.1124 to 0.2929) | <0.0001 |
| **CD81** | 0.6195 (0.5319 to 0.7216) | <0.0001 | 0.0227 | 0.2605 (0.1640 to 0.3571) | <0.0001 |
| CD9 | 1.4831 (1.2776 to 1.7216) | <0.0001 | 0.0160 | -0.2146 (-0.3100 to -0.1191) | <0.0001 |
| **HLA-ABC** | 1.5791 (1.3548 to 1.8406) | <0.0001 | 0.0202 | -0.2075 (-0.3021 to -0.1128) | <0.0001 |
| HLA-DR/ DP/DQ | 0.6950 (0.5987 to 0.8068) | <0.0001 | 0.0136 | 0.2082 (0.1123 to 0.3041) | <0.0001 |

**Table S3b.3: Cross-sectional associations of EV markers with COPD severity: sensitivity analysis with additional adjustment for FVC z-score (full GOLD 0–4 cohort, prespecified covariates + FVC z-score). OR**: odds ratio for higher GOLD stage per standard-deviation marker increase (ordinal logistic regression). **β**: coefficient for $FEV_1$ z-score per SD (linear regression, HC3-robust SE). **p**: raw p-value. **$R^2$**: McFadden's pseudo-$R^2$ for the ordinal model (left block) and the linear regression's $R^2$ for the right block. **$\Delta R^2$**: incremental marker contribution. **$pR^2$**: marker contribution to the linear regression. Significance is reported as raw p-values; FDR adjustment is applied only in the primary analysis (Table S3b.1). **Bold**: top-tier markers (CD8, CD29, CD31, CD49e, CD81, HLA-ABC; see Table S3a).

| Marker | **Ordinal logistic regression** (GOLD stages) | | | | | **Linear regression** (FEV1 z-score) | | |
|---|---|---|---|---|---|---|---|---|
| | **OR** (CI 95%) | **p** | **$R^2$** | **$\Delta R^2$** | **LR p** | **β** (95% CI) | **p** | **$pR^2$** |
| CD105 | 1.2031 (1.0164 to 1.4241) | 0.0316 | 0.2699 | 0.0029 | 0.0312 | -0.0403 (-0.0998 to 0.0191) | 0.1838 | 0.0011 |
| CD142 | 1.1073 (0.9392 to 1.3055) | 0.2251 | 0.2679 | 0.0009 | 0.2248 | -0.0106 (-0.0670 to 0.0458) | 0.7134 | 0.0001 |
| CD146 | 1.2653 (1.0696 to 1.4967) | 0.0060 | 0.2717 | 0.0048 | 0.0058 | -0.0690 (-0.1311 to -0.0068) | 0.0296 | 0.0032 |
| CD24 | 0.9028 (0.7627 to 1.0685) | 0.2341 | 0.2678 | 0.0009 | 0.2338 | 0.0000 (-0.0624 to 0.0625) | 0.9988 | 0.0000 |
| CD25 | 0.9250 (0.7840 to 1.0914) | 0.3558 | 0.2675 | 0.0005 | 0.3559 | 0.0203 (-0.0385 to 0.0791) | 0.4989 | 0.0003 |
| **CD29** | 1.4914 (1.2510 to 1.7780) | <0.0001 | 0.2797 | 0.0127 | <0.0001 | -0.1105 (-0.1673 to -0.0537) | 0.0001 | 0.0082 |
| CD3 | 0.9339 (0.7940 to 1.0985) | 0.4089 | 0.2674 | 0.0004 | 0.4087 | 0.0175 (-0.0390 to 0.0741) | 0.5436 | 0.0002 |
| **CD31** | 1.3591 (1.1482 to 1.6088) | 0.0004 | 0.2751 | 0.0081 | 0.0003 | -0.0958 (-0.1521 to -0.0395) | 0.0008 | 0.0062 |
| CD40 | 1.1045 (0.9348 to 1.3050) | 0.2430 | 0.2678 | 0.0009 | 0.2428 | -0.0349 (-0.0940 to 0.0241) | 0.2461 | 0.0008 |
| CD42a | 1.0952 (0.9315 to 1.2876) | 0.2708 | 0.2677 | 0.0008 | 0.2712 | -0.0268 (-0.0820 to 0.0283) | 0.3400 | 0.0005 |
| CD44 | 0.8272 (0.6995 to 0.9783) | 0.0267 | 0.2700 | 0.0031 | 0.0262 | 0.0496 (-0.0088 to 0.1080) | 0.0960 | 0.0016 |
| CD45 | 0.9660 (0.8197 to 1.1383) | 0.6794 | 0.2670 | 0.0001 | 0.6795 | 0.0056 (-0.0526 to 0.0637) | 0.8514 | 0.0000 |
| **CD49e** | 1.3400 (1.1276 to 1.5925) | 0.0009 | 0.2740 | 0.0070 | 0.0008 | -0.0994 (-0.1565 to -0.0424) | 0.0006 | 0.0065 |

| **Marker** | **Ordinal logistic regression** (GOLD stages) | | | | | **Linear regression** (FEV1 z-score) | | |
|---|---|---|---|---|---|---|---|---|
| | **OR** (CI 95%) | **p** | **$R^2$** | **$\Delta R^2$** | **LR p** | **β** (95% CI) | **p** | **$pR^2$** |
| CD56 | 1.0283 (0.8746 to 1.2091) | 0.7353 | 0.2670 | 0.0001 | 0.7353 | -0.0155 (-0.0689 to 0.0379) | 0.5692 | 0.0002 |
| CD62P | 0.9559 (0.8108 to 1.1270) | 0.5913 | 0.2671 | 0.0002 | 0.5910 | 0.0077 (-0.0478 to 0.0633) | 0.7854 | 0.0000 |
| CD63 | 0.9826 (0.8351 to 1.1562) | 0.8326 | 0.2670 | 0.0000 | 0.8326 | 0.0284 (-0.0276 to 0.0845) | 0.3196 | 0.0005 |
| CD69 | 1.3317 (1.1242 to 1.5776) | 0.0009 | 0.2739 | 0.0070 | 0.0008 | -0.0855 (-0.1433 to -0.0277) | 0.0037 | 0.0049 |
| **CD8** | 0.7706 (0.6520 to 0.9107) | 0.0022 | 0.2729 | 0.0060 | 0.0021 | 0.0898 (0.0384 to 0.1413) | 0.0006 | 0.0054 |
| **CD81** | 0.7381 (0.6208 to 0.8776) | 0.0006 | 0.2745 | 0.0075 | 0.0005 | 0.0977 (0.0370 to 0.1583) | 0.0016 | 0.0062 |
| CD9 | 1.3057 (1.1049 to 1.5430) | 0.0017 | 0.2731 | 0.0062 | 0.0017 | -0.0876 (-0.1463 to -0.0290) | 0.0034 | 0.0051 |
| **HLA-ABC** | 1.4963 (1.2559 to 1.7828) | <0.0001 | 0.2800 | 0.0131 | <0.0001 | -0.1045 (-0.1613 to -0.0478) | 0.0003 | 0.0072 |
| HLA-DR/DP/DQ | 0.8430 (0.7122 to 0.9977) | 0.0470 | 0.2694 | 0.0025 | 0.0464 | 0.0734 (0.0188 to 0.1280) | 0.0085 | 0.0034 |

**Table S3b.4: Cross-sectional associations of EV markers with COPD severity: sensitivity analysis restricted to established COPD (GOLD 1–4, prespecified covariates). OR**: odds ratio for higher GOLD stage per standard-deviation marker increase (ordinal logistic regression). **β**: coefficient for $FEV_1$ z-score per SD (linear regression, HC3-robust SE). **p**: raw p-value. **$R^2$**: McFadden's pseudo-$R^2$ for the ordinal model (left block) and the linear regression's $R^2$ for the right block. **$\Delta R^2$**: incremental marker contribution. **$pR^2$**: marker contribution to the linear regression. Significance is reported as raw p-values; FDR adjustment is applied only in the primary analysis (Table S3b.1). **Bold**: top-tier markers (CD8, CD29, CD31, CD49e, CD81, HLA-ABC; see Table S3a).

| **Marker** | **Ordinal logistic regression** (GOLD stages) | | | | | **Linear regression** ($FEV_1$ z-score) | | |
|---|---|---|---|---|---|---|---|---|
| | **OR** (CI 95%) | **p** | **$R^2$** | **$\Delta R^2$** | **LR p** | **β** (95% CI) | **p** | **$pR^2$** |
| CD105 | 0.9068 (0.7641 to 1.0761) | 0.2624 | 0.1734 | 0.0010 | 0.2620 | 0.0657 (-0.0168 to 0.1483) | 0.1184 | 0.0031 |
| CD142 | 0.9547 (0.8024 to 1.1360) | 0.6016 | 0.1727 | 0.0002 | 0.6013 | 0.0453 (-0.0437 to 0.1342) | 0.3187 | 0.0014 |
| CD146 | 1.2194 (1.0289 to 1.4451) | 0.0221 | 0.1766 | 0.0041 | 0.0214 | -0.0822 (-0.1637 to -0.0007) | 0.0480 | 0.0051 |
| CD24 | 0.9330 (0.7849 to 1.1090) | 0.4316 | 0.1729 | 0.0005 | 0.4314 | 0.0160 (-0.0756 to 0.1076) | 0.7316 | 0.0002 |
| CD25 | 0.9563 (0.8075 to 1.1325) | 0.6044 | 0.1727 | 0.0002 | 0.6043 | 0.0283 (-0.0558 to 0.1123) | 0.5095 | 0.0006 |
| **CD29** | 1.2854 (1.0848 to 1.5231) | 0.0037 | 0.1791 | 0.0067 | 0.0035 | -0.1070 (-0.1834 to -0.0306) | 0.0061 | 0.0086 |
| CD3 | 1.0588 (0.8973 to 1.2493) | 0.4986 | 0.1728 | 0.0004 | 0.4983 | -0.0186 (-0.0964 to 0.0592) | 0.6391 | 0.0003 |
| **CD31** | 1.2587 (1.0620 to 1.4918) | 0.0079 | 0.1780 | 0.0056 | 0.0076 | -0.0919 (-0.1720 to -0.0118) | 0.0246 | 0.0063 |
| CD40 | 1.0705 (0.9033 to 1.2686) | 0.4317 | 0.1729 | 0.0005 | 0.4314 | -0.0104 (-0.0936 to 0.0728) | 0.8064 | 0.0001 |
| CD42a | 1.1717 (0.9921 to 1.3839) | 0.0620 | 0.1752 | 0.0027 | 0.0617 | -0.0578 (-0.1359 to 0.0203) | 0.1470 | 0.0025 |
| CD44 | 0.9007 (0.7562 to 1.0727) | 0.2408 | 0.1735 | 0.0011 | 0.2408 | 0.0368 (-0.0555 to 0.1292) | 0.4341 | 0.0010 |
| CD45 | 1.0376 (0.8759 to 1.2292) | 0.6695 | 0.1726 | 0.0001 | 0.6693 | -0.0059 (-0.0901 to 0.0782) | 0.8901 | 0.0000 |
| **CD49e** | 1.2231 (1.0307 to 1.4515) | 0.0211 | 0.1766 | 0.0042 | 0.0207 | -0.0908 (-0.1689 to -0.0126) | 0.0229 | 0.0060 |

| **Marker** | **Ordinal logistic regression** (GOLD stages) | | | | | **Linear regression** ($FEV_1$ z-score) | | |
|---|---|---|---|---|---|---|---|---|
| | **OR** (CI 95%) | **p** | **$R^2$** | **$\Delta R^2$** | **LR p** | **β** (95% CI) | **p** | **p$R^2$** |
| CD56 | 1.0481 (0.8889 to 1.2358) | 0.5763 | 0.1727 | 0.0002 | 0.5763 | -0.0056 (-0.0808 to 0.0695) | 0.8830 | 0.0000 |
| CD62P | 0.9634 (0.8092 to 1.1469) | 0.6750 | 0.1726 | 0.0001 | 0.6746 | 0.0095 (-0.0788 to 0.0978) | 0.8336 | 0.0001 |
| CD63 | 0.9395 (0.7922 to 1.1141) | 0.4730 | 0.1729 | 0.0004 | 0.4730 | 0.0547 (-0.0308 to 0.1402) | 0.2100 | 0.0022 |
| CD69 | 1.1758 (0.9937 to 1.3912) | 0.0592 | 0.1753 | 0.0028 | 0.0587 | -0.0708 (-0.1494 to 0.0078) | 0.0774 | 0.0037 |
| **CD8** | 0.8378 (0.7072 to 0.9926) | 0.0408 | 0.1758 | 0.0033 | 0.0403 | 0.0912 (0.0129 to 0.1696) | 0.0225 | 0.0062 |
| **CD81** | 0.7937 (0.6670 to 0.9446) | 0.0093 | 0.1778 | 0.0053 | 0.0090 | 0.1006 (0.0168 to 0.1845) | 0.0187 | 0.0072 |
| CD9 | 1.1840 (1.0006 to 1.4010) | 0.0491 | 0.1755 | 0.0030 | 0.0487 | -0.0744 (-0.1555 to 0.0067) | 0.0720 | 0.0041 |
| **HLA-ABC** | 1.2421 (1.0478 to 1.4723) | 0.0125 | 0.1774 | 0.0049 | 0.0121 | -0.0804 (-0.1589 to -0.0019) | 0.0448 | 0.0048 |
| HLA-DR/ DP/DQ | 0.8906 (0.7484 to 1.0598) | 0.1916 | 0.1738 | 0.0013 | 0.1911 | 0.0767 (-0.0085 to 0.1620) | 0.0778 | 0.0042 |

**Table S3b.5: Cross-sectional associations of EV markers with COPD severity: sensitivity analysis without covariates, restricted to established COPD (GOLD 1–4). OR**: odds ratio for higher GOLD stage per standard-deviation marker increase (ordinal logistic regression). **β**: coefficient for $FEV_1$ z-score per SD (linear regression, HC3-robust SE). **p**: raw p-value. **$R^2$**: McFadden's pseudo-$R^2$ for the ordinal model. Significance is reported as raw p-values; FDR adjustment is applied only in the primary analysis (Table S3b.1). **Bold**: top-tier markers (CD8, CD29, CD31, CD49e, CD81, HLA-ABC; see Table S3a).

| **Marker** | **Ordinal logistic regression** (GOLD stages) | | | **Linear regression** (FEV1 z-score) | |
|---|---|---|---|---|---|
| | **OR** (CI 95%) | **p** | **$R^2$** | **β** (95% CI) | **p** |
| CD105 | 0.8839 (0.7556 to 1.0339) | 0.1228 | 0.0018 | 0.0846 (-0.0137 to 0.1828) | 0.0917 |
| CD142 | 0.9136 (0.7792 to 1.0712) | 0.2659 | 0.0009 | 0.0572 (-0.0455 to 0.1599) | 0.2747 |
| CD146 | 1.2218 (1.0397 to 1.4359) | 0.0150 | 0.0044 | -0.1073 (-0.2097 to -0.0048) | 0.0402 |
| CD24 | 0.9359 (0.8009 to 1.0936) | 0.4042 | 0.0005 | 0.0327 (-0.0694 to 0.1348) | 0.5306 |
| CD25 | 0.9311 (0.7972 to 1.0875) | 0.3677 | 0.0006 | 0.0426 (-0.0547 to 0.1398) | 0.3911 |
| **CD29** | 1.3163 (1.1213 to 1.5452) | 0.0008 | 0.0085 | -0.1526 (-0.2517 to -0.0536) | 0.0025 |
| CD3 | 1.0315 (0.8839 to 1.2036) | 0.6940 | 0.0001 | -0.0240 (-0.1174 to 0.0693) | 0.6139 |
| **CD31** | 1.2999 (1.1084 to 1.5245) | 0.0013 | 0.0078 | -0.1360 (-0.2331 to -0.0390) | 0.0060 |
| CD40 | 1.1326 (0.9688 to 1.3241) | 0.1182 | 0.0018 | -0.0651 (-0.1635 to 0.0333) | 0.1949 |
| CD42a | 1.0532 (0.8988 to 1.2340) | 0.5216 | 0.0003 | -0.0329 (-0.1319 to 0.0661) | 0.5149 |
| CD44 | 0.8242 (0.7029 to 0.9663) | 0.0172 | 0.0042 | 0.0949 (-0.0093 to 0.1992) | 0.0743 |
| CD45 | 0.9843 (0.8415 to 1.1514) | 0.8434 | 0.0000 | 0.0076 (-0.0920 to 0.1071) | 0.8813 |
| **CD49e** | 1.2320 (1.0514 to 1.4437) | 0.0099 | 0.0049 | -0.1196 (-0.2175 to -0.0216) | 0.0167 |
| CD56 | 0.9883 (0.8472 to 1.1530) | 0.8815 | 0.0000 | 0.0116 (-0.0825 to 0.1058) | 0.8086 |

| **Marker** | **Ordinal logistic regression** (GOLD stages) | | | **Linear regression** (FEV1 z-score) | |
|---|---|---|---|---|---|
| | **OR** (CI 95%) | **p** | **$R^2$** | **β** (95% CI) | **p** |
| CD62P | 0.9847 (0.8404 to 1.1537) | 0.8485 | 0.0000 | 0.0047 (-0.0987 to 0.1080) | 0.9297 |
| CD63 | 0.9341 (0.7977 to 1.0937) | 0.3970 | 0.0005 | 0.0619 (-0.0386 to 0.1624) | 0.2273 |
| CD69 | 1.2756 (1.0908 to 1.4916) | 0.0023 | 0.0069 | -0.1357 (-0.2321 to -0.0392) | 0.0058 |
| **CD8** | 0.8090 (0.6918 to 0.9460) | 0.0079 | 0.0053 | 0.1250 (0.0331 to 0.2170) | 0.0077 |
| **CD81** | 0.7334 (0.6261 to 0.8591) | 0.0001 | 0.0110 | 0.1709 (0.0736 to 0.2683) | 0.0006 |
| CD9 | 1.2386 (1.0601 to 1.4472) | 0.0070 | 0.0054 | -0.1241 (-0.2199 to -0.0282) | 0.0112 |
| **HLA-ABC** | 1.2263 (1.0465 to 1.4371) | 0.0117 | 0.0047 | -0.1011 (-0.1990 to -0.0033) | 0.0428 |
| HLA-DR/DP/DQ | 0.7652 (0.6536 to 0.8959) | 0.0009 | 0.0083 | 0.1620 (0.0630 to 0.2610) | 0.0013 |

**Table S3b.6: Cross-sectional associations of EV markers with COPD severity: sensitivity analysis with additional adjustment for RV/TLC z-score, restricted to established COPD (GOLD 1–4, prespecified covariates + RV/TLC z-score). OR**: odds ratio for higher GOLD stage per standard-deviation marker increase (ordinal logistic regression). **β**: coefficient for $FEV_1$ z-score per SD (linear regression, HC3-robust SE). **p**: raw p-value. **$R^2$**: McFadden's pseudo-$R^2$ for the ordinal model (left block) and the linear regression's $R^2$ for the right block. **$\Delta R^2$**: incremental marker contribution. **$pR^2$**: marker contribution to the linear regression. Significance is reported as raw p-values; FDR adjustment is applied only in the primary analysis (Table S3b.1). **Bold**: top-tier markers (CD8, CD29, CD31, CD49e, CD81, HLA-ABC; see Table S3a).

| **Marker** | **Ordinal logistic regression** (GOLD stages) | | | | | **Linear regression** ($FEV_1$ z-score) | | |
|---|---|---|---|---|---|---|---|---|
| | **OR** (CI 95%) | **p** | **$R^2$** | **$\Delta R^2$** | **LR p** | **β** (95% CI) | **p** | **$pR^2$** |
| CD105 | 0.9304 (0.7791 to 1.1110) | 0.4254 | 0.1715 | 0.0005 | 0.4247 | 0.0574 (-0.0288 to 0.1437) | 0.1920 | 0.0023 |
| CD142 | 0.9468 (0.7929 to 1.1305) | 0.5454 | 0.1713 | 0.0003 | 0.5459 | 0.0487 (-0.0414 to 0.1388) | 0.2892 | 0.0017 |
| CD146 | 1.2005 (1.0096 to 1.4276) | 0.0386 | 0.1745 | 0.0036 | 0.0379 | -0.0772 (-0.1606 to 0.0062) | 0.0698 | 0.0045 |
| CD24 | 0.9265 (0.7761 to 1.1061) | 0.3983 | 0.1716 | 0.0006 | 0.4025 | 0.0142 (-0.0796 to 0.1081) | 0.7662 | 0.0001 |
| CD25 | 0.9402 (0.7904 to 1.1184) | 0.4860 | 0.1714 | 0.0004 | 0.4861 | 0.0339 (-0.0535 to 0.1214) | 0.4469 | 0.0008 |
| CD29 | 1.2907 (1.0862 to 1.5337) | 0.0037 | 0.1780 | 0.0071 | 0.0035 | -0.1079 (-0.1846 to -0.0313) | 0.0058 | 0.0088 |
| CD3 | 1.0499 (0.8857 to 1.2446) | 0.5747 | 0.1712 | 0.0003 | 0.5803 | -0.0183 (-0.0993 to 0.0627) | 0.6576 | 0.0002 |
| CD31 | 1.2553 (1.0566 to 1.4914) | 0.0097 | 0.1766 | 0.0056 | 0.0094 | -0.0916 (-0.1718 to -0.0115) | 0.0250 | 0.0063 |
| CD40 | 1.0466 (0.8805 to 1.2441) | 0.6054 | 0.1712 | 0.0002 | 0.6067 | -0.0007 (-0.0855 to 0.0840) | 0.9864 | 0.0000 |
| CD42a | 1.1766 (0.9929 to 1.3942) | 0.0604 | 0.1739 | 0.0029 | 0.0602 | -0.0627 (-0.1418 to 0.0164) | 0.1204 | 0.0030 |
| CD44 | 0.8902 (0.7436 to 1.0657) | 0.2052 | 0.1723 | 0.0013 | 0.2051 | 0.0384 (-0.0571 to 0.1340) | 0.4302 | 0.0010 |
| CD45 | 1.0230 (0.8581 to 1.2197) | 0.7994 | 0.1710 | 0.0001 | 0.8000 | -0.0047 (-0.0929 to 0.0836) | 0.9173 | 0.0000 |
| CD49e | 1.2173 (1.0220 to 1.4499) | 0.0275 | 0.1750 | 0.0040 | 0.0270 | -0.0900 (-0.1700 to -0.0100) | 0.0275 | 0.0059 |

| **Marker** | **Ordinal logistic regression** (GOLD stages) | | | | | **Linear regression** ($FEV_1$ z-score) | | |
|---|---|---|---|---|---|---|---|---|
| | **OR** (CI 95%) | **p** | **R²** | **ΔR²** | **LR p** | **β** (95% CI) | **p** | **pR²** |
| CD56 | 1.0429 (0.8823 to 1.2329) | 0.6224 | 0.1712 | 0.0002 | 0.6224 | -0.0070 (-0.0825 to 0.0684) | 0.8550 | 0.0000 |
| CD62P | 0.9594 (0.8008 to 1.1494) | 0.6532 | 0.1711 | 0.0002 | 0.6534 | 0.0145 (-0.0775 to 0.1064) | 0.7579 | 0.0001 |
| CD63 | 0.9414 (0.7900 to 1.1217) | 0.4994 | 0.1714 | 0.0004 | 0.5005 | 0.0602 (-0.0276 to 0.1480) | 0.1788 | 0.0027 |
| CD69 | 1.1544 (0.9723 to 1.3707) | 0.1013 | 0.1732 | 0.0022 | 0.1008 | -0.0657 (-0.1458 to 0.0144) | 0.1080 | 0.0032 |
| **CD8** | 0.8298 (0.6950 to 0.9907) | 0.0391 | 0.1745 | 0.0035 | 0.0386 | 0.0871 (0.0044 to 0.1698) | 0.0389 | 0.0054 |
| CD81 | 0.7941 (0.6647 to 0.9486) | 0.0110 | 0.1763 | 0.0054 | 0.0108 | 0.1037 (0.0194 to 0.1879) | 0.0159 | 0.0077 |
| CD9 | 1.1678 (0.9835 to 1.3868) | 0.0767 | 0.1736 | 0.0026 | 0.0762 | -0.0695 (-0.1525 to 0.0135) | 0.1009 | 0.0036 |
| **HLA-ABC** | 1.2417 (1.0437 to 1.4772) | 0.0146 | 0.1760 | 0.0050 | 0.0142 | -0.0836 (-0.1634 to -0.0038) | 0.0400 | 0.0052 |
| HLA-DR/DP/DQ | 0.8930 (0.7463 to 1.0685) | 0.2163 | 0.1722 | 0.0013 | 0.2162 | 0.0656 (-0.0225 to 0.1537) | 0.1445 | 0.0030 |

**Table S4: Jonckheere-Terpstra trend tests for monotonic association between EV marker levels and GOLD stage.** Two-sided tests across ordered GOLD groups in the full cohort (GOLD 0–4) and restricted to GOLD 1–4. **z**: standardised test statistic (positive = marker levels increase with GOLD stage). **p**: raw p-value. **q**: Benjamini-Hochberg-adjusted p across 22 markers in the primary (full-cohort) analysis; raw p-values only in the GOLD 1–4 sensitivity analysis. **Bold**: top-tier markers (CD8, CD29, CD31, CD49e, CD81, HLA-ABC; see Table S3a).

| **Marker** | **Full GOLD 0–4 cohort** | | | **GOLD 1–4 cohort** | |
|---|---|---|---|---|---|
| | **z** | **q** | **p** | **z** | **p** |
| CD105 | 0.1585 | 0.8741 | 0.8741 | -1.4108 | 0.1583 |
| CD142 | -0.9776 | 0.4514 | 0.3283 | -1.3152 | 0.1884 |
| CD146 | 3.0346 | 0.0048 | 0.0024 | 1.5134 | 0.1302 |
| CD24 | -1.9768 | 0.0813 | 0.0481 | -0.8094 | 0.4183 |
| CD25 | -1.6983 | 0.1406 | 0.0894 | -1.1071 | 0.2683 |
| **CD29** | 4.7671 | <0.0001 | <0.0001 | 2.7415 | 0.0061 |
| CD3 | -0.8307 | 0.4703 | 0.4062 | -0.0784 | 0.9375 |
| **CD31** | 3.2645 | 0.0024 | 0.0011 | 1.8159 | 0.0694 |
| CD40 | 2.0508 | 0.0739 | 0.0403 | 1.4098 | 0.1586 |
| CD42a | -0.6769 | 0.5483 | 0.4985 | -0.1979 | 0.8431 |
| CD44 | -4.3131 | <0.0001 | <0.0001 | -2.3208 | 0.0203 |
| CD45 | -0.3386 | 0.7699 | 0.7349 | -0.2833 | 0.7770 |
| **CD49e** | 4.1063 | <0.0001 | <0.0001 | 1.9182 | 0.0551 |
| CD56 | -0.9179 | 0.4641 | 0.3587 | -0.8141 | 0.4156 |
| CD62P | -0.8435 | 0.4703 | 0.3990 | -0.2196 | 0.8262 |
| CD63 | -1.4969 | 0.1972 | 0.1344 | -0.4248 | 0.6710 |
| CD69 | 4.5815 | <0.0001 | <0.0001 | 2.6270 | 0.0086 |
| **CD8** | -5.1572 | <0.0001 | <0.0001 | -2.7732 | 0.0056 |
| **CD81** | -5.9624 | <0.0001 | <0.0001 | -3.6854 | 0.0002 |
| CD9 | 4.3439 | <0.0001 | <0.0001 | 1.8743 | 0.0609 |
| **HLA-ABC** | 4.1428 | <0.0001 | <0.0001 | 1.4660 | 0.1427 |
| HLA-DR/DP/DQ | -4.6403 | <0.0001 | <0.0001 | -3.0932 | 0.0020 |

**Table S5: Mann-Whitney U tests of baseline EV marker levels between progressors and non-progressors.** Progression: ≥1 GOLD-stage increase over 54 months. Tests performed in the full cohort (GOLD 0–4) and restricted to GOLD 1–4. **U**: Mann-Whitney U statistic (two-sided). **p**: raw p-value. **q**: Benjamini-Hochberg-adjusted p across 22 markers in the primary (full-cohort) analysis; raw p-values only in the GOLD 1–4 sensitivity analysis. **Bold**: markers below q < 0.05.

| **Marker** | **Full GOLD 0–4 cohort** | | | **GOLD 1–4 cohort** | |
|---|---|---|---|---|---|
| | **U** | **q** | **p** | **U** | **p** |
| **CD105** | 83884 | <0.0001 | <0.0001 | 71340 | <0.0001 |
| **CD142** | 88735 | <0.0001 | <0.0001 | 75255 | <0.0001 |
| CD146 | 69202 | 0.6698 | 0.6090 | 57420 | 0.2246 |
| **CD24** | 88945 | <0.0001 | <0.0001 | 75255 | <0.0001 |
| **CD25** | 81235 | 0.0010 | 0.0003 | 68121 | 0.0033 |
| CD29 | 69490 | 0.7124 | 0.6801 | 56985 | 0.1666 |
| **CD3** | 77207 | 0.0389 | 0.0248 | 63945 | 0.1883 |
| CD31 | 69971 | 0.8051 | 0.8051 | 57507 | 0.2377 |
| **CD40** | 79284 | 0.0062 | 0.0031 | 66642 | 0.0182 |
| **CD42a** | 77505 | 0.0320 | 0.0189 | 63771 | 0.2120 |
| CD44 | 76533 | 0.0607 | 0.0441 | 64206 | 0.1565 |
| **CD45** | 76848 | 0.0497 | 0.0339 | 63858 | 0.1999 |
| CD49e | 74863 | 0.1948 | 0.1505 | 63423 | 0.2656 |
| **CD56** | 80371 | 0.0024 | 0.0009 | 66381 | 0.0238 |
| **CD62P** | 78996 | 0.0077 | 0.0042 | 66207 | 0.0283 |
| **CD63** | 82878 | 0.0001 | <0.0001 | 70383 | 0.0001 |
| **CD69** | 79643 | 0.0045 | 0.0020 | 68295 | 0.0027 |
| **CD8** | 79836 | 0.0040 | 0.0016 | 66033 | 0.0336 |
| **CD81** | 81994 | 0.0004 | <0.0001 | 68904 | 0.0012 |
| CD9 | 74592 | 0.2186 | 0.1789 | 62727 | 0.3991 |
| HLA-ABC | 73799 | 0.3289 | 0.2841 | 59682 | 0.7360 |
| **HLA-DR/DP/DQ** | 81025 | 0.0012 | 0.0004 | 67947 | 0.0041 |

**Table S6a: Longitudinal associations of baseline EV markers with $FEV_1$ trajectory: primary analysis (full GOLD 0–4 cohort).** Linear mixed-effects models with repeated $FEV_1$ z-score measurements as the dependent variable. Each marker was modelled separately with subject-specific random intercepts and random slopes for follow-up time (years from baseline, centred at the cohort mean), adjusted for the prespecified covariate set (Methods). **β (marker)**: fixed-effect coefficient for the standardised baseline marker, representing the cross-sectional association at the centring time. **β (time×marker)**: fixed-effect coefficient for the marker-by-time interaction, representing the per-year modification of $FEV_1$ trajectory per standard-deviation marker increase. **p**: raw p-value for the corresponding fixed effect. **q**: Benjamini-Hochberg-adjusted p across 22 markers, computed separately for the main effect and the interaction term. **$R^2$m**: marginal $R^2$ (variance explained by fixed effects alone; Nakagawa & Schielzeth, 2013). **$R^2$c**: conditional $R^2$ (variance explained by fixed and random effects combined). All models converged.

| Marker | Marker | | | Time × marker | | | $R^2$m | $R^2$c |
|---|---|---|---|---|---|---|---|---|
| | β | p | q | β | p | q | | |
| CD105 | 0.1150 | 0.0024 | 0.0044 | -0.0057 | 0.3808 | 0.5647 | 0.1198 | 0.8996 |
| CD142 | 0.0970 | 0.0112 | 0.0163 | 0.0007 | 0.9112 | 0.9546 | 0.1161 | 0.9001 |
| CD146 | -0.1119 | 0.0099 | 0.0156 | 0.0065 | 0.2201 | 0.5233 | 0.1016 | 0.9077 |
| CD24 | 0.0800 | 0.0597 | 0.0657 | 0.0095 | 0.0741 | 0.5233 | 0.0957 | 0.9073 |
| CD25 | 0.1397 | 0.0008 | 0.0015 | 0.0062 | 0.2408 | 0.5233 | 0.1073 | 0.9088 |
| CD29 | -0.1758 | <0.0001 | <0.0001 | 0.0065 | 0.2735 | 0.5233 | 0.1381 | 0.8987 |
| CD3 | 0.0926 | 0.0125 | 0.0163 | 0.0079 | 0.1949 | 0.5233 | 0.1180 | 0.9028 |
| CD31 | -0.1609 | <0.0001 | <0.0001 | 0.0082 | 0.1683 | 0.5233 | 0.1341 | 0.8992 |
| CD40 | -0.1477 | 0.0002 | 0.0006 | 0.0044 | 0.3850 | 0.5647 | 0.1143 | 0.9080 |
| CD42a | -0.0576 | 0.1743 | 0.1826 | 0.0002 | 0.9681 | 0.9681 | 0.0922 | 0.9071 |
| CD44 | 0.1604 | 0.0001 | 0.0004 | 0.0008 | 0.8808 | 0.9546 | 0.1154 | 0.9086 |
| CD45 | 0.0777 | 0.0393 | 0.0480 | 0.0084 | 0.1782 | 0.5233 | 0.1145 | 0.9033 |

| Marker | Marker | | | Time × marker | | | $R^2m$ | $R^2c$ |
|---|---|---|---|---|---|---|---|---|
| | **β** | **p** | **q** | **β** | **p** | **q** | | |
| CD49e | -0.1339 | 0.0006 | 0.0014 | 0.0059 | 0.3319 | 0.5616 | 0.1257 | 0.8990 |
| CD56 | 0.1126 | 0.0073 | 0.0123 | 0.0081 | 0.1320 | 0.5233 | 0.1020 | 0.9084 |
| CD62P | -0.1407 | 0.0007 | 0.0015 | 0.0023 | 0.6593 | 0.9065 | 0.1107 | 0.9073 |
| CD63 | -0.0113 | 0.7678 | 0.7678 | 0.0019 | 0.7685 | 0.9546 | 0.1087 | 0.9001 |
| CD69 | -0.1081 | 0.0126 | 0.0163 | 0.0080 | 0.1413 | 0.5233 | 0.1015 | 0.9078 |
| CD8 | 0.2078 | <0.0001 | <0.0001 | -0.0012 | 0.8177 | 0.9546 | 0.1343 | 0.9085 |
| **CD81** | 0.2387 | <0.0001 | <0.0001 | -0.0090 | 0.0888 | 0.5233 | 0.1452 | 0.9092 |
| CD9 | -0.1629 | <0.0001 | <0.0001 | 0.0064 | 0.2854 | 0.5233 | 0.1332 | 0.8998 |
| HLA-ABC | -0.0914 | 0.0442 | 0.0512 | 0.0082 | 0.1581 | 0.5233 | 0.0966 | 0.9073 |
| HLA-DR/DP/DQ | 0.2052 | <0.0001 | <0.0001 | 0.0008 | 0.8847 | 0.9546 | 0.1515 | 0.8995 |

**Table S6b: Longitudinal associations of baseline EV markers with $FEV_1$ trajectory: sensitivity analysis restricted to established COPD (GOLD 1–4).** Linear mixed-effects models as in Table S6a. **β (marker)**: fixed-effect coefficient for the standardised baseline marker. **β (time×marker)**: fixed-effect coefficient for the marker-by-time interaction. **p**: raw p-value for the corresponding fixed effect. **$R^2$m**: marginal $R^2$ (fixed effects only). **$R^2$c**: conditional $R^2$ (fixed + random effects). Significance is reported as raw p-values; FDR adjustment is applied only in the primary analysis (Table S6a). All models converged.

| **Marker** | **Marker** | | **Time × marker** | | **$R^2$m** | **$R^2$c** |
|---|---|---|---|---|---|---|
| | **β** | **p** | **β** | **p** | | |
| CD105 | 0.0925 | 0.0159 | -0.0044 | 0.4955 | 0.1336 | 0.8982 |
| CD142 | 0.0999 | 0.0195 | 0.0018 | 0.7451 | 0.1178 | 0.9076 |
| CD146 | -0.0426 | 0.2739 | 0.0051 | 0.3989 | 0.1276 | 0.8972 |
| CD24 | 0.0490 | 0.2030 | 0.0113 | 0.0699 | 0.1285 | 0.8990 |
| CD25 | 0.0609 | 0.1545 | 0.0090 | 0.1056 | 0.1120 | 0.9073 |
| CD29 | -0.0698 | 0.0805 | 0.0058 | 0.3687 | 0.1302 | 0.8975 |
| CD3 | 0.0183 | 0.6346 | 0.0126 | 0.0414 | 0.1258 | 0.9006 |
| CD31 | -0.0450 | 0.2606 | 0.0088 | 0.1694 | 0.1277 | 0.8977 |
| CD40 | 0.0147 | 0.7296 | 0.0035 | 0.5434 | 0.1082 | 0.9069 |
| CD42a | 0.0130 | 0.7380 | 0.0001 | 0.9876 | 0.1257 | 0.8973 |
| CD44 | 0.0782 | 0.0668 | 0.0022 | 0.6933 | 0.1159 | 0.9075 |
| CD45 | 0.0356 | 0.3567 | 0.0092 | 0.1319 | 0.1268 | 0.8993 |
| CD49e | -0.0437 | 0.2651 | 0.0066 | 0.2874 | 0.1276 | 0.8973 |
| CD56 | 0.0375 | 0.3338 | 0.0129 | 0.0421 | 0.1268 | 0.9001 |
| CD62P | -0.0014 | 0.9744 | -0.0027 | 0.6602 | 0.1083 | 0.9069 |
| CD63 | -0.0067 | 0.8641 | -0.0033 | 0.6040 | 0.1253 | 0.8971 |
| CD69 | -0.0670 | 0.1148 | 0.0110 | 0.0503 | 0.1143 | 0.9073 |
| CD8 | 0.1109 | 0.0042 | -0.0025 | 0.6816 | 0.1408 | 0.8990 |
| **CD81** | 0.0993 | 0.0204 | -0.0123 | 0.0332 | 0.1197 | 0.9075 |
| CD9 | -0.0289 | 0.4693 | 0.0082 | 0.1946 | 0.1265 | 0.8978 |
| HLA-ABC | -0.0158 | 0.7220 | 0.0088 | 0.1564 | 0.1088 | 0.9070 |
| HLA-DR/DP/DQ | 0.1190 | 0.0022 | 0.0004 | 0.9508 | 0.1414 | 0.8999 |

**Table S7a: Cox proportional hazards regression for EV markers and all-cause mortality: primary analysis (Core + $FEV_1$z covariates).** Cox models fitted with the prespecified core covariate set (sex, age, leukocyte count, $PaCO_2$, platelets), $FEV_1$ z-score and Efron handling of tied event times; robust sandwich standard errors. Each marker was modelled separately. **HR**: hazard ratio per standard-deviation increase in standardised baseline marker level. **95% CI**: Wald 95% confidence interval. **p**: raw p-value. **q**: Benjamini-Hochberg-adjusted p across 22 markers. **Schoenfeld p**: scaled-Schoenfeld-residual test of the proportional hazards assumption (significant values suggest non-proportional hazards). **Bold**: top-tier prognostic markers (CD25, CD56, CD142; significant Cox association across all adjustment schemes). Sensitivity analyses with raw p-values: Tables S7b-S7g.

| Marker | HR | 95% CI | p | q | Schoenfeld p |
|---|---|---|---|---|---|
| CD105 | 0.816 | 0.681-0.978 | 0.0277 | 0.1180 | 0.2831 |
| **CD142** | 0.740 | 0.605-0.904 | 0.0033 | 0.0241 | 0.5086 |
| CD146 | 1.024 | 0.870-1.205 | 0.7783 | 0.8154 | 0.6666 |
| CD24 | 0.808 | 0.665-0.982 | 0.0322 | 0.1180 | 0.9939 |
| **CD25** | 0.766 | 0.649-0.904 | 0.0016 | 0.0177 | 0.5541 |
| CD29 | 0.963 | 0.802-1.156 | 0.6826 | 0.7904 | 0.7907 |
| CD3 | 0.869 | 0.740-1.022 | 0.0890 | 0.2447 | 0.3921 |
| CD31 | 0.913 | 0.745-1.119 | 0.3820 | 0.6139 | 0.5775 |
| CD40 | 0.914 | 0.746-1.119 | 0.3832 | 0.6139 | 0.0704 |
| CD42a | 0.927 | 0.762-1.127 | 0.4465 | 0.6139 | 0.8953 |
| CD44 | 0.944 | 0.776-1.148 | 0.5607 | 0.7016 | 0.0157 |
| CD45 | 0.927 | 0.776-1.108 | 0.4061 | 0.6139 | 0.0613 |
| CD49e | 0.886 | 0.738-1.063 | 0.1924 | 0.6139 | 0.1209 |
| ***CD56*** | 0.749 | 0.627-0.894 | 0.0014 | 0.0177 | 0.8551 |
| CD62P | 0.858 | 0.675-1.092 | 0.2143 | 0.4715 | 0.0919 |
| CD63 | 1.071 | 0.903-1.270 | 0.4331 | 0.6139 | 0.0199 |
| CD69 | 0.946 | 0.780-1.148 | 0.5741 | 0.7016 | 0.0274 |
| CD8 | 1.1 | 0.900-1.344 | 0.3529 | 0.6139 | 0.0009 |
| CD81 | 1.003 | 0.787-1.278 | 0.9809 | 0.9809 | 0.0062 |
| CD9 | 1.035 | 0.848-1.264 | 0.7347 | 0.8083 | 0.3825 |
| HLA-ABC | 0.804 | 0.659-0.981 | 0.0318 | 0.1180 | 0.6121 |
| HLA-DR/DP/DQ | 0.819 | 0.673-0.997 | 0.0471 | 0.1479 | 0.1258 |

T**able S7b: Cox proportional hazards regression for EV markers and all-cause mortality: sensitivity analysis with only clinical covariates (Core).** Cox models as in Table S7a. **HR**: hazard ratio per standard-deviation increase in standardised baseline marker level. **95% CI**: Wald 95% confidence interval. **p**: raw p-value. **Schoenfeld p**: scaled-Schoenfeld-residual test of the proportional hazards assumption. Significance is reported as raw p-values; FDR adjustment is applied only in the primary analysis (Table S7a). **Bold**: top-tier prognostic markers (CD25, CD56, CD142; see Table S7a).

| **Marker** | **HR** | **95% CI** | **p** | **Schoenfeld p** |
|---|---|---|---|---|
| CD105 | 0.824 | 0.682-0.994 | 0.0432 | 0.8735 |
| **CD142** | 0.695 | 0.566-0.853 | 0.0005 | 0.1525 |
| CD146 | 0.985 | 0.835-1.161 | 0.8543 | 0.4674 |
| CD24 | 0.813 | 0.678-0.974 | 0.025 | 0.6281 |
| **CD25** | 0.732 | 0.624-0.860 | 0.0001 | 0.4386 |
| CD29 | 0.987 | 0.836-1.164 | 0.8734 | 0.5459 |
| CD3 | 0.826 | 0.703-0.970 | 0.0194 | 0.5817 |
| CD31 | 0.942 | 0.791-1.123 | 0.5066 | 0.3751 |
| CD40 | 0.926 | 0.761-1.126 | 0.4393 | 0.0661 |
| CD42a | 0.893 | 0.735-1.084 | 0.2515 | 0.4709 |
| CD44 | 0.839 | 0.690-1.019 | 0.0771 | 0.0062 |
| CD45 | 0.82 | 0.698-0.964 | 0.0159 | 0.1297 |
| CD49e | 0.897 | 0.748-1.076 | 0.2417 | 0.1048 |
| ***CD56*** | 0.740 | 0.620-0.884 | 0.0009 | 0.8293 |
| CD62P | 0.846 | 0.681-1.052 | 0.1335 | 0.028 |
| CD63 | 1.068 | 0.884-1.290 | 0.4958 | 0.0775 |
| CD69 | 1.02 | 0.850-1.224 | 0.8312 | 0.0235 |
| CD8 | 0.989 | 0.816-1.197 | 0.9075 | 0.0006 |
| CD81 | 0.95 | 0.748-1.207 | 0.6761 | 0.0085 |
| CD9 | 1.015 | 0.846-1.218 | 0.8697 | 0.1471 |
| HLA-ABC | 0.826 | 0.687-0.992 | 0.0404 | 0.3666 |
| HLA-DR/DP/DQ | 0.743 | 0.609-0.908 | 0.0036 | 0.0721 |

**Table S7c: Cox proportional hazards regression for EV markers and all-cause mortality: sensitivity analysis with additional adjustment for FVC z-score (Core + FVCz).** Cox models as in Table S7a. **HR**: hazard ratio per standard-deviation increase in standardised baseline marker level. **95% CI**: Wald 95% confidence interval. **p**: raw p-value. **Schoenfeld p**: scaled-Schoenfeld-residual test of the proportional hazards assumption. Significance is reported as raw p-values; FDR adjustment is applied only in the primary analysis (Table S7a). **Bold**: top-tier prognostic markers (CD25, CD56, CD142; see Table S7a).

| Marker | HR | 95% CI | p | Schoenfeld p |
|---|---|---|---|---|
| CD105 | 0.857 | 0.715-1.027 | 0.0940 | 0.3030 |
| **CD142** | 0.754 | 0.616-0.923 | 0.0061 | 0.4113 |
| CD146 | 1.016 | 0.856-1.205 | 0.8583 | 0.7786 |
| CD24 | 0.818 | 0.675-0.990 | 0.039 | 0.8303 |
| **CD25** | 0.755 | 0.631-0.905 | 0.0024 | 0.3293 |
| CD29 | 1.016 | 0.852-1.212 | 0.8572 | 0.6817 |
| CD3 | 0.851 | 0.707-1.024 | 0.087 | 0.2284 |
| CD31 | 0.97 | 0.794-1.186 | 0.7684 | 0.5312 |
| CD40 | 0.937 | 0.768-1.142 | 0.5169 | 0.0742 |
| CD42a | 0.931 | 0.767-1.131 | 0.4738 | 0.5783 |
| CD44 | 0.904 | 0.739-1.106 | 0.3257 | 0.0048 |
| CD45 | 0.889 | 0.736-1.072 | 0.2178 | 0.0299 |
| CD49e | 0.936 | 0.769-1.138 | 0.5064 | 0.1868 |
| ***CD56*** | 0.768 | 0.628-0.939 | 0.0100 | 0.6702 |
| CD62P | 0.853 | 0.678-1.073 | 0.1741 | 0.0497 |
| CD63 | 1.064 | 0.893-1.268 | 0.4886 | 0.0296 |
| CD69 | 1.018 | 0.843-1.229 | 0.8530 | 0.0275 |
| CD8 | 1.026 | 0.838-1.256 | 0.8040 | 0.0002 |
| CD81 | 0.955 | 0.754-1.211 | 0.7043 | 0.0018 |
| CD9 | 1.052 | 0.872-1.270 | 0.5942 | 0.2640 |
| HLA-ABC | 0.851 | 0.698-1.038 | 0.1118 | 0.5797 |
| HLA-DR/DP/DQ | 0.769 | 0.631-0.938 | 0.0096 | 0.0309 |

**Table S7d: Cox proportional hazards regression for EV markers and all-cause mortality: sensitivity analysis with additional adjustment for RV/TLC z-score (Core + RV/TLCz).** Cox models as in Table S7a. **HR**: hazard ratio per standard-deviation increase in standardised baseline marker level. **95% CI**: Wald 95% confidence interval. **p**: raw p-value. **Schoenfeld p**: scaled-Schoenfeld-residual test of the proportional hazards assumption. Significance is reported as raw p-values; FDR adjustment is applied only in the primary analysis (Table S7a). **Bold**: top-tier prognostic markers (CD25, CD56, CD142; see Table S7a).

| Marker | HR | 95% CI | p | Schoenfeld p |
|---|---|---|---|---|
| CD105 | 0.818 | 0.661-1.013 | 0.0651 | 0.7455 |
| **CD142** | 0.691 | 0.555-0.862 | 0.001 | 0.1655 |
| CD146 | 0.939 | 0.792-1.113 | 0.4682 | 0.3947 |
| CD24 | 0.829 | 0.684-1.004 | 0.0552 | 0.4137 |
| **CD25** | 0.719 | 0.607-0.851 | 0.0001 | 0.7504 |
| CD29 | 0.935 | 0.787-1.110 | 0.4432 | 0.5180 |
| CD3 | 0.816 | 0.696-0.958 | 0.0131 | 0.9118 |
| CD31 | 0.899 | 0.750-1.077 | 0.2482 | 0.3058 |
| CD40 | 0.887 | 0.711-1.107 | 0.2894 | 0.0656 |
| CD42a | 0.849 | 0.696-1.034 | 0.1032 | 0.6428 |
| CD44 | 0.879 | 0.708-1.092 | 0.2444 | 0.0145 |
| CD45 | 0.834 | 0.708-0.983 | 0.0306 | 0.1752 |
| CD49e | 0.866 | 0.714-1.049 | 0.1416 | 0.0829 |
| ***CD56*** | 0.713 | 0.595-0.854 | 0.0002 | 0.9755 |
| CD62P | 0.843 | 0.668-1.065 | 0.1531 | 0.0053 |
| CD63 | 1.119 | 0.921-1.360 | 0.2585 | 0.0631 |
| CD69 | 0.983 | 0.801-1.205 | 0.8671 | 0.0347 |
| CD8 | 1.076 | 0.854-1.355 | 0.5327 | 0.0039 |
| CD81 | 1.009 | 0.777-1.310 | 0.9462 | 0.0132 |
| CD9 | 0.968 | 0.801-1.169 | 0.7325 | 0.1808 |
| HLA-ABC | 0.79 | 0.651-0.959 | 0.0169 | 0.2223 |
| HLA-DR/DP/DQ | 0.768 | 0.612-0.964 | 0.0229 | 0.0943 |

**Table S7e: Cox proportional hazards regression for EV markers and all-cause mortality: sensitivity analysis with additional adjustment for $FEV_1$ z-score and FVC z-score (Core + $FEV_1$z + FVCz).** Cox models as in Table S7a. **HR**: hazard ratio per standard-deviation increase in standardised baseline marker level. **95% CI**: Wald 95% confidence interval. **p**: raw p-value. **Schoenfeld p**: scaled-Schoenfeld-residual test of the proportional hazards assumption. Significance is reported as raw p-values; FDR adjustment is applied only in the primary analysis (Table S7a). **Bold**: top-tier prognostic markers (CD25, CD56, CD142; see Table S7a).

| Marker | HR | 95% CI | p | Schoenfeld p |
|---|---|---|---|---|
| CD105 | 0.824 | 0.686-0.989 | 0.0375 | 0.2830 |
| **CD142** | 0.744 | 0.607-0.911 | 0.0043 | 0.5097 |
| CD146 | 1.023 | 0.867-1.206 | 0.7882 | 0.7333 |
| CD24 | 0.81 | 0.666-0.986 | 0.0353 | 0.9864 |
| **CD25** | 0.765 | 0.645-0.907 | 0.002 | 0.5035 |
| CD29 | 0.976 | 0.812-1.175 | 0.8008 | 0.8629 |
| CD3 | 0.866 | 0.732-1.025 | 0.0947 | 0.3263 |
| CD31 | 0.926 | 0.752-1.141 | 0.4709 | 0.650 |
| CD40 | 0.919 | 0.750-1.126 | 0.4155 | 0.0779 |
| CD42a | 0.932 | 0.767-1.133 | 0.4802 | 0.8158 |
| CD44 | 0.944 | 0.775-1.151 | 0.5704 | 0.0122 |
| CD45 | 0.925 | 0.772-1.110 | 0.4037 | 0.0477 |
| CD49e | 0.897 | 0.743-1.082 | 0.2557 | 0.1543 |
| ***CD56*** | 0.752 | 0.628-0.901 | 0.002 | 0.7895 |
| CD62P | 0.859 | 0.676-1.092 | 0.2153 | 0.0921 |
| CD63 | 1.072 | 0.904-1.272 | 0.4241 | 0.0213 |
| CD69 | 0.958 | 0.787-1.166 | 0.6703 | 0.0353 |
| CD8 | 1.094 | 0.893-1.340 | 0.3877 | 0.0006 |
| CD81 | 0.998 | 0.784-1.269 | 0.9847 | 0.0046 |
| CD9 | 1.039 | 0.852-1.268 | 0.7030 | 0.4082 |
| HLA-ABC | 0.812 | 0.664-0.992 | 0.0410 | 0.6758 |
| HLA-DR/DP/DQ | 0.81 | 0.665-0.988 | 0.0372 | 0.0946 |

**Table S7f: Cox proportional hazards regression for EV markers and all-cause mortality: sensitivity analysis with additional adjustment for $FEV_1$ z-score and RV/TLC z-score (Core + $FEV_1$z + RV/TLCz).** Cox models as in Table S7a. **HR**: hazard ratio per standard-deviation increase in standardised baseline marker level. **95% CI**: Wald 95% confidence interval. **p**: raw p-value. **Schoenfeld p**: scaled-Schoenfeld-residual test of the proportional hazards assumption. Significance is reported as raw p-values; FDR adjustment is applied only in the primary analysis (Table S7a). **Bold**: top-tier prognostic markers (CD25, CD56, CD142; see Table S7a).

| Marker | HR | 95% CI | p | Schoenfeld p |
|---|---|---|---|---|
| CD105 | 0.802 | 0.657-0.980 | 0.0309 | 0.2711 |
| **CD142** | 0.746 | 0.604-0.921 | 0.0064 | 0.4465 |
| CD146 | 1.011 | 0.846-1.208 | 0.9039 | 0.3802 |
| CD24 | 0.825 | 0.674-1.010 | 0.0625 | 0.8682 |
| **CD25** | 0.765 | 0.643-0.911 | 0.0026 | 0.846 |
| CD29 | 0.931 | 0.767-1.129 | 0.4676 | 0.5864 |
| CD3 | 0.875 | 0.742-1.032 | 0.1134 | 0.6681 |
| CD31 | 0.889 | 0.715-1.106 | 0.2909 | 0.4035 |
| CD40 | 0.892 | 0.709-1.122 | 0.3281 | 0.0677 |
| CD42a | 0.893 | 0.725-1.100 | 0.2863 | 0.7706 |
| CD44 | 0.969 | 0.786-1.194 | 0.7650 | 0.0331 |
| CD45 | 0.952 | 0.796-1.139 | 0.5918 | 0.0856 |
| CD49e | 0.865 | 0.714-1.047 | 0.1368 | 0.1011 |
| ***CD56*** | 0.729 | 0.607-0.877 | 0.0008 | 0.9725 |
| CD62P | 0.843 | 0.655-1.084 | 0.1830 | 0.0224 |
| CD63 | 1.108 | 0.925-1.326 | 0.2649 | 0.0078 |
| CD69 | 0.929 | 0.748-1.153 | 0.5028 | 0.0378 |
| CD8 | 1.196 | 0.953-1.501 | 0.1230 | 0.0061 |
| CD81 | 1.038 | 0.798-1.352 | 0.7795 | 0.0060 |
| CD9 | 1.026 | 0.830-1.269 | 0.8096 | 0.2310 |
| HLA-ABC | 0.793 | 0.640-0.982 | 0.0331 | 0.3144 |
| HLA-DR/DP/DQ | 0.837 | 0.669-1.045 | 0.1164 | 0.1031 |

**Table S7g: Cox proportional hazards regression for EV markers and all-cause mortality: sensitivity analysis with additional adjustment for FVC z-score and RV/TLC z-score (Core + FVCz + RV/TLCz).** Cox models as in Table S7a. **HR**: hazard ratio per standard-deviation increase in standardised baseline marker level. **95% CI**: Wald 95% confidence interval. **p**: raw p-value. **Schoenfeld p**: scaled-Schoenfeld-residual test of the proportional hazards assumption. Significance is reported as raw p-values; FDR adjustment is applied only in the primary analysis (Table S7a). **Bold**: top-tier prognostic markers (CD25, CD56, CD142; see Table S7a).

| **Marker** | **HR** | **95% CI** | **p** | **Schoenfeld p** |
|---|---|---|---|---|
| CD105 | 0.848 | 0.695-1.034 | 0.1039 | 0.3240 |
| **CD142** | 0.765 | 0.617-0.948 | 0.0146 | 0.4232 |
| CD146 | 1.001 | 0.830-1.207 | 0.9899 | 0.5223 |
| CD24 | 0.829 | 0.676-1.018 | 0.0729 | 0.7028 |
| **CD25** | 0.758 | 0.626-0.916 | 0.0042 | 0.5653 |
| CD29 | 0.986 | 0.817-1.190 | 0.8793 | 0.5874 |
| CD3 | 0.861 | 0.715-1.036 | 0.1134 | 0.4443 |
| CD31 | 0.948 | 0.763-1.177 | 0.6290 | 0.4279 |
| CD40 | 0.918 | 0.732-1.150 | 0.4564 | 0.0923 |
| CD42a | 0.901 | 0.732-1.110 | 0.3275 | 0.8595 |
| CD44 | 0.935 | 0.754-1.159 | 0.5406 | 0.0129 |
| CD45 | 0.92 | 0.763-1.110 | 0.3859 | 0.062 |
| CD49e | 0.919 | 0.747-1.131 | 0.4270 | 0.1541 |
| ***CD56*** | 0.752 | 0.613-0.923 | 0.0063 | 0.8235 |
| CD62P | 0.837 | 0.660-1.062 | 0.1439 | 0.0135 |
| CD63 | 1.097 | 0.913-1.319 | 0.3213 | 0.0147 |
| CD69 | 1.005 | 0.814-1.240 | 0.9647 | 0.0501 |
| CD8 | 1.119 | 0.890-1.406 | 0.3367 | 0.0016 |
| CD81 | 0.995 | 0.767-1.289 | 0.9674 | 0.0024 |
| CD9 | 1.039 | 0.848-1.273 | 0.7152 | 0.1883 |
| HLA-ABC | 0.843 | 0.680-1.045 | 0.1185 | 0.3169 |
| HLA-DR/DP/DQ | 0.788 | 0.628-0.989 | 0.0401 | 0.0348 |

**Table S8a: Incremental prognostic performance of CD25 across all lung function adjustment schemes.** Reclassification at the 54-month horizon. **Adjustment**: lung function variables added to the Core covariate set (sex, age, leukocytes, $PaCO_2$, platelets). **C**: Harrell's C-index of the extended model (Core + lung function + CD25). **ΔC**: difference vs the reference model without CD25. **cNRI**: continuous net reclassification improvement. **catNRI**: categorical NRI (cut-points 5%, 10%, 20%). **IDI**: integrated discrimination improvement. **n**: participants with known event status at horizon. 95% CI and two-sided bootstrap p (5000 resamples) reported alongside each estimate. Parallel results for CD56, CD142 and CD25 + CD69: Tables S8b-S8d.

| Adjustment | C | ΔC | | | cNRI | | | catNRI | | | IDI | | | n |
|---|---|---|---|---|---|---|---|---|---|---|---|---|---|---|
| | | ΔC | 95% CI | p | cNRI | 95% CI | p | catNRI | 95% CI | p | IDI | 95% CI | p | |
| Core | 0.7917 | 0.0235 | 0.0011 to 0.0473 | 0.0404 | 0.3784 | 0.0876 to 0.6625 | 0.0124 | 0.0937 | -0.0500 to 0.2373 | 0.1952 | 0.0211 | 0.0053 to 0.0373 | 0.0088 | 281 |
| Core + $FEV_1z$ | 0.8323 | 0.0090 | -0.0033 to 0.0223 | 0.1560 | 0.3519 | 0.0555 to 0.6360 | 0.0204 | 0.1828 | 0.0577 to 0.3105 | 0.0056 | 0.0169 | 0.0019 to 0.0319 | 0.0260 | 281 |
| Core + FVCz | 0.8184 | 0.0131 | -0.0016 to 0.0296 | 0.0840 | 0.4590 | 0.1719 to 0.7281 | 0.0040 | 0.1068 | -0.0209 to 0.2365 | 0.0940 | 0.0197 | 0.0045 to 0.0347 | 0.0100 | 281 |
| Core + RV/TLCz | 0.7798 | 0.0264 | -0.0017 to 0.0549 | 0.0632 | 0.3668 | 0.0433 to 0.6827 | 0.0264 | 0.1334 | -0.0140 to 0.2826 | 0.0772 | 0.0230 | 0.0067 to 0.0405 | 0.0044 | 266 |
| Core + FEV1z + FVCz | 0.8332 | 0.0100 | -0.0024 to 0.0237 | 0.1152 | 0.3519 | 0.0555 to 0.6360 | 0.0204 | 0.1604 | 0.0472 to 0.2827 | 0.0072 | 0.0177 | 0.0028 to 0.0326 | 0.0184 | 281 |
| Core + $FEV_1z$ + RV/TLCz | 0.8194 | 0.0094 | -0.0057 to 0.0251 | 0.2388 | 0.2830 | -0.0322 to 0.6015 | 0.0748 | 0.1668 | 0.0378 to 0.2981 | 0.0076 | 0.0174 | 0.0022 to 0.0344 | 0.0284 | 266 |
| Core + FVCz + RV/TLCz | 0.8065 | 0.0136 | -0.0037 to 0.0326 | 0.1356 | 0.2395 | -0.0726 to 0.5545 | 0.1332 | 0.0227 | -0.1160 to 0.1569 | 0.7528 | 0.0191 | 0.0041 to 0.0352 | 0.0112 | 266 |

**Table S8b: Incremental prognostic performance of CD56 across all lung function adjustment schemes.** Reclassification at the 54-month horizon. **Adjustment**: lung function variables added to the Core covariate set (sex, age, leukocytes, $PaCO_2$, platelets). **C**: Harrell's C-index of the extended model (Core + lung function + CD56). **ΔC**: difference vs the reference model without CD56. **cNRI**: continuous net reclassification improvement. **catNRI**: categorical NRI (cut-points 5%, 10%, 20%). **IDI**: integrated discrimination improvement. **n**: participants with known event status at horizon. 95% CI and two-sided bootstrap p (5000 resamples) reported alongside each estimate. Parallel results for CD25, CD142 and CD25 + CD69: Tables S8a, S8c, S8d.

| **Adjustment** | **C** | **ΔC** | | | **cNRI** | | | **catNRI** | | | **IDI** | | | **n** |
|---|---|---|---|---|---|---|---|---|---|---|---|---|---|---|
| | | **ΔC** | **95% CI** | **p** | **cNRI** | **95% CI** | **p** | **catNRI** | **95% CI** | **p** | **IDI** | **95% CI** | **p** | |
| Core | 0.7891 | 0.0210 | -0.0003 to 0.0441 | 0.0552 | 0.3070 | 0.0107 to 0.5884 | 0.0428 | 0.1337 | -0.0083 to 0.2777 | 0.0640 | 0.0177 | 0.0025 to 0.0340 | 0.0220 | 281 |
| Core + $FEV_1z$ | 0.8370 | 0.0137 | -0.0004 to 0.0307 | 0.0616 | 0.2359 | -0.0613 to 0.5243 | 0.1212 | 0.1159 | -0.0088 to 0.2405 | 0.0656 | 0.0134 | -0.0030 to 0.0299 | 0.1064 | 281 |
| Core + FVCz | 0.8175 | 0.0122 | -0.0023 to 0.0297 | 0.1080 | 0.2451 | -0.0529 to 0.5265 | 0.1028 | 0.1248 | 0.0000 to 0.2521 | 0.0516 | 0.0135 | -0.0014 to 0.0286 | 0.0744 | 281 |
| Core + RV/TLCz | 0.7813 | 0.0279 | 0.0014 to 0.0573 | 0.0372 | 0.3486 | 0.0238 to 0.6589 | 0.0328 | 0.1208 | -0.0354 to 0.2734 | 0.1308 | 0.0236 | 0.0068 to 0.0406 | 0.0040 | 266 |
| Core + $FEV_1z$ + FVCz | 0.8369 | 0.0137 | 0.0000 to 0.0303 | 0.0504 | 0.2537 | -0.0483 to 0.5420 | 0.0928 | 0.0892 | -0.0059 to 0.1907 | 0.0724 | 0.0135 | -0.0027 to 0.0300 | 0.1016 | 281 |
| Core + $FEV_1z$ + RV/TLCz | 0.8256 | 0.0156 | -0.0023 to 0.0378 | 0.0972 | 0.1273 | -0.2042 to 0.4373 | 0.4600 | 0.0798 | -0.0568 to 0.2182 | 0.2500 | 0.0178 | -0.0008 to 0.0357 | 0.0580 | 266 |
| Core + FVCz + RV/TLCz | 0.8088 | 0.0159 | -0.0016 to 0.0367 | 0.0792 | 0.1597 | -0.1736 to 0.4696 | 0.3432 | 0.1233 | 0.0121 to 0.2428 | 0.0340 | 0.0160 | 0.0002 to 0.0317 | 0.0464 | 266 |

**Table S8c: Incremental prognostic performance of CD142 across all lung function adjustment schemes.** Reclassification at the 54-month horizon. **Adjustment**: lung function variables added to the Core covariate set (sex, age, leukocytes, $PaCO_2$, platelets). **C**: Harrell's C-index of the extended model (Core + lung function + CD142). **ΔC**: difference vs the reference model without CD142. **cNRI**: continuous net reclassification improvement. **catNRI**: categorical NRI (cut-points 5%, 10%, 20%). **IDI**: integrated discrimination improvement. **n**: participants with known event status at horizon. 95% CI and two-sided bootstrap p (5000 resamples) reported alongside each estimate. Parallel results for CD25, CD56 and CD25 + CD69: Tables S8a, S8b, S8d.

| Adjustment | C | ΔC | | | cNRI | | | catNRI | | | IDI | | | n |
|---|---|---|---|---|---|---|---|---|---|---|---|---|---|---|
| | | ΔC | 95% CI | p | cNRI | 95% CI | p | catNRI | 95% CI | p | IDI | 95% CI | p | |
| Core | 0.7820 | 0.0139 | -0.0137 to 0.0414 | 0.3016 | 0.5840 | 0.3058 to 0.8648 | <0.0001 | 0.1114 | -0.0258 to 0.2585 | 0.1204 | 0.0377 | 0.0199 to 0.0562 | <0.0001 | 281 |
| Core + $FEV_1z$ | 0.8336 | 0.0103 | -0.0032 to 0.0247 | 0.1284 | 0.4414 | 0.1592 to 0.7229 | 0.0028 | 0.1560 | 0.0251 to 0.2949 | 0.0188 | 0.0204 | 0.0042 to 0.0359 | 0.0152 | 281 |
| Core + FVCz | 0.8168 | 0.0115 | -0.0034 to 0.0273 | 0.1372 | 0.3344 | 0.0415 to 0.6231 | 0.0288 | 0.1513 | 0.0238 to 0.2913 | 0.0216 | 0.0151 | 0.0015 to 0.0286 | 0.0272 | 281 |
| Core + RV/TLCz | 0.7669 | 0.0135 | -0.0205 to 0.0478 | 0.4392 | 0.5206 | 0.2131 to 0.8091 | 0.0012 | 0.0889 | -0.0752 to 0.2498 | 0.2976 | 0.0363 | 0.0173 to 0.0558 | <0.0001 | 266 |
| Core + $FEV_1z$ + FVCz | 0.8358 | 0.0126 | -0.0005 to 0.0268 | 0.0612 | 0.3344 | 0.0409 to 0.6259 | 0.0284 | 0.1293 | 0.0048 to 0.2612 | 0.0432 | 0.0183 | 0.0026 to 0.0335 | 0.0216 | 281 |
| Core + $FEV_1z$ + RV/TLCz | 0.8194 | 0.0093 | -0.0049 to 0.0241 | 0.2076 | 0.3862 | 0.0783 to 0.6682 | 0.0148 | 0.0925 | -0.0637 to 0.2408 | 0.2512 | 0.0174 | 0.0001 to 0.0328 | 0.0484 | 266 |
| Core + FVCz + RV/TLCz | 0.8041 | 0.0112 | -0.0051 to 0.0274 | 0.1860 | 0.3862 | 0.0765 to 0.6739 | 0.0124 | 0.1117 | -0.0390 to 0.2561 | 0.1416 | 0.0111 | -0.0027 to 0.0236 | 0.1084 | 266 |

**Table S8d: Incremental prognostic performance of the CD25 + CD69 combination across all lung function adjustment schemes.** Reclassification at the 54-month horizon. CD25 + CD69 was identified by the predefined stability-based selection procedure. **Adjustment**: lung function variables added to the Core covariate set (sex, age, leukocytes, $PaCO_2$, platelets). **C**: Harrell's C-index of the extended model (Core + lung function + CD25 + CD69). **ΔC**: difference vs the reference model without the EV markers. **cNRI**: continuous net reclassification improvement. **catNRI**: categorical NRI (cut-points 5%, 10%, 20%). **IDI**: integrated discrimination improvement. **n**: participants with known event status at horizon. 95% CI and two-sided bootstrap p (5000 resamples) reported alongside each estimate. Single-marker results: Tables S8a (CD25), S8b (CD56), S8c (CD142).

| Adjustment | C | ΔC | | | cNRI | | | catNRI | | | IDI | | | n |
|---|---|---|---|---|---|---|---|---|---|---|---|---|---|---|
| | | ΔC | 95% CI | p | cNRI | 95% CI | p | catNRI | 95% CI | p | IDI | 95% CI | p | |
| Core | 0.8138 | 0.0457 | 0.0185 to 0.0734 | 0.0008 | 0.5483 | 0.2738 to 0.8167 | 0.0004 | 0.2675 | 0.0957 to 0.4352 | 0.0016 | 0.0370 | 0.0111 to 0.0620 | 0.0040 | 281 |
| Core + $FEV_1z$ | 0.8401 | 0.0168 | 0.0020 to 0.0325 | 0.0268 | 0.4679 | 0.1821 to 0.7339 | 0.0016 | 0.1828 | 0.0455 to 0.3236 | 0.0112 | 0.0238 | 0.0045 to 0.0433 | 0.0192 | 281 |
| Core + FVCz | 0.8333 | 0.0280 | 0.0077 to 0.0502 | 0.0048 | 0.5305 | 0.2487 to 0.8058 | 0.0004 | 0.2718 | 0.1345 to 0.4125 | <0.0001 | 0.0367 | 0.0114 to 0.0607 | 0.0044 | 281 |
| Core + RV/TLCz | 0.8000 | 0.0466 | 0.0131 to 0.0811 | 0.0060 | 0.5822 | 0.2785 to 0.8709 | 0.0004 | 0.2441 | 0.0702 to 0.4234 | 0.0056 | 0.0378 | 0.0123 to 0.0640 | 0.0032 | 266 |
| Core + $FEV_1z$ + FVCz | 0.8420 | 0.0188 | 0.0028 to 0.0358 | 0.0228 | 0.5394 | 0.2599 to 0.8093 | 0.0004 | 0.1648 | 0.0263 to 0.3048 | 0.0220 | 0.0270 | 0.0065 to 0.0473 | 0.0100 | 281 |
| Core + $FEV_1z$ + RV/TLCz | 0.8275 | 0.0175 | 0.0004 to 0.0351 | 0.0432 | 0.3609 | 0.0513 to 0.6643 | 0.0236 | 0.1324 | -0.0244 to 0.2913 | 0.1068 | 0.0232 | 0.0029 to 0.0452 | 0.0228 | 266 |
| Core + FVCz + RV/TLCz | 0.8219 | 0.0290 | 0.0049 to 0.0547 | 0.0116 | 0.4933 | 0.1829 to 0.7878 | 0.0008 | 0.2340 | 0.0819 to 0.3817 | 0.0040 | 0.0355 | 0.0090 to 0.0623 | 0.0068 | 266 |

**Table S9a: Single marker selection for combined models.** Stability: marker selection stability from repeated nested cross-validated elastic-net Cox regression without any covariates. Coefficient: EV markers by absolute standardized coefficient from the final ridge-penalized Cox model adjusted for clinical covariates. **Uns. coefficient**: unsigned coefficients. **Bold**: the five most stable markers, which are also markers with the highest unsigned coefficients.

| **Marker** | **Stability** | **Coefficient** | **Uns. coefficient** |
|---|---|---|---|
| CD105 | 0.8500 | -0.0835 | 0.0835 |
| **CD142** | 0.9800 | -0.1195 | 0.1195 |
| CD146 | 0.3600 | 0.0256 | 0.0256 |
| CD24 | 0.3100 | -0.0166 | 0.0166 |
| **CD25** | 0.9200 | -0.1015 | 0.1015 |
| CD29 | 0.6100 | 0.0379 | 0.0379 |
| CD3 | 0.3400 | 0.0228 | 0.0228 |
| CD31 | 0.1700 | -0.0030 | 0.0030 |
| CD40 | 0.3600 | 0.0040 | 0.0040 |
| CD42a | 0.2700 | -0.0145 | 0.0145 |
| CD44 | 0.3000 | -0.0362 | 0.0362 |
| CD45 | 0.2700 | -0.0155 | 0.0155 |
| CD49e | 0.1800 | -0.0037 | 0.0037 |
| CD56 | 0.8900 | -0.0982 | 0.0982 |
| **CD62P** | 0.9900 | -0.1357 | 0.1357 |
| CD63 | 0.8900 | 0.0895 | 0.0895 |
| **CD69** | 0.9400 | 0.1031 | 0.1031 |
| CD8 | 0.2600 | 0.0303 | 0.0303 |
| CD81 | 0.6000 | -0.0446 | 0.0446 |
| CD9 | 0.2800 | 0.0206 | 0.0206 |
| **HLA-ABC** | 0.9300 | -0.1292 | 0.1292 |
| HLA-DR/DP/DQ | 0.6300 | -0.0837 | 0.0837 |

**Table S9b: Combined markers.** Incremental prognostic value of all pairwise combinations drawn from the five highest-ranked EV markers (**Table S9a**) added to the clinical backbone Cox model. Discrimination gain (ΔC-index with bootstrap 95% CI and two-sided p-value), reclassification (continuous and categorical NRI), and integrated discrimination improvement (IDI) are shown for each combination, sorted by ΔC-index. **Bold**: select combination for further analyses.

| Markers | C-index | ΔC-index | ΔC-index 95% CI | p | Cont. NRI | Cat. NRI | IDI |
|---|---|---|---|---|---|---|---|
| **CD25 + CD69** | 0.840 | 0.017 | 0.002 to 0.032 | 0.0268 | 0.468 | 0.183 | 0.024 |
| CD62P + CD142 | 0.839 | 0.015 | 0.002 to 0.029 | 0.0200 | 0.441 | 0.174 | 0.022 |
| CD142 + CD25 | 0.838 | 0.014 | 0.000 to 0.031 | 0.0580 | 0.53 | 0.196 | 0.027 |
| CD142 + CD69 | 0.834 | 0.011 | -0.003 to 0.025 | 0.1088 | 0.397 | 0.138 | 0.021 |
| CD62P + CD25 | 0.834 | 0.01 | -0.003 to 0.024 | 0.1132 | 0.406 | 0.174 | 0.019 |
| HLA-ABC + CD142 | 0.832 | 0.009 | -0.007 to 0.025 | 0.2812 | 0.593 | 0.053 | 0.025 |
| HLA-ABC + CD25 | 0.831 | 0.007 | -0.004 to 0.021 | 0.2224 | 0.459 | 0.178 | 0.017 |
| CD62P + HLA-ABC | 0.829 | 0.006 | -0.008 to 0.022 | 0.4112 | 0.263 | 0.134 | 0.013 |
| CD62P + CD69 | 0.827 | 0.004 | -0.005 to 0.012 | 0.3632 | 0.093 | 0.08 | 0.005 |
| HLA-ABC + CD69 | 0.825 | 0.002 | -0.011 to 0.016 | 0.7828 | 0.316 | 0.111 | 0.013 |

**Table S9c: Cross-validated optimism correction for the four primary prognostic configurations.** Repeated stratified 10-fold cross-validation (100 repeats; 1000 evaluations per configuration) of the reference model (Core + $FEV_1z$) and each extended model. **Apparent**: full-data estimate (Table 3). **CV-median**: median across the 1000 held-out evaluations. **CV percentile (95%)**: fold-to-fold variability, reported as a stability diagnostic rather than a confidence interval. **Optimism**: apparent - CV-median; values near zero indicate the apparent estimate is not materially inflated by in-sample fitting. Reclassification metrics computed on participants with known event status at the 54-month horizon; categorical NRI cut-points 5%, 10%, 20%.

| Configuration | Metric | Apparent | CV-median | CV percentile (95%) | Optimism |
|---|---|---|---|---|---|
| CD25 | C-index | 0.832 | 0.838 | 0.635 to 0.950 | -0.006 |
| | ΔC-index | 0.009 | 0.008 | -0.035 to 0.054 | 0.001 |
| | Continuous NRI | 0.352 | 0.369 | -0.560 to 1.286 | -0.017 |
| | Categorical NRI | 0.183 | 0.105 | -0.248 to 0.536 | 0.078 |
| | IDI | 0.017 | 0.017 | -0.032 to 0.069 | 0.000 |
| CD56 | C-index | 0.837 | 0.845 | 0.636 to 0.948 | -0.008 |
| | ΔC-index | 0.014 | 0.01 | -0.032 to 0.069 | 0.004 |
| | Continuous NRI | 0.236 | 0.226 | -0.648 to 1.217 | 0.010 |
| | Categorical NRI | 0.116 | 0.056 | -0.298 to 0.444 | 0.060 |
| | IDI | 0.013 | 0.014 | -0.045 to 0.059 | -0.001 |
| CD142 | C-index | 0.834 | 0.838 | 0.631 to 0.949 | -0.004 |
| | ΔC-index | 0.01 | 0.01 | -0.043 to 0.059 | -0.000 |
| | Continuous NRI | 0.441 | 0.386 | -0.543 to 1.263 | 0.055 |
| | Categorical NRI | 0.156 | 0.058 | -0.333 to 0.511 | 0.098 |
| | IDI | 0.02 | 0.021 | -0.038 to 0.065 | -0.001 |
| CD25 + CD69 | C-index | 0.84 | 0.845 | 0.639 to 0.954 | -0.005 |
| | ΔC-index | 0.017 | 0.013 | -0.035 to 0.071 | 0.004 |
| | Continuous NRI | 0.468 | 0.441 | -0.515 to 1.300 | 0.027 |
| | Categorical NRI | 0.183 | 0.12 | -0.250 to 0.567 | 0.063 |
| | IDI | 0.024 | 0.021 | -0.046 to 0.089 | 0.003 |
